# Interpretable machine learning prediction of all-cause mortality

**DOI:** 10.1101/2021.01.20.21250135

**Authors:** Wei Qiu, Hugh Chen, Ayse Berceste Dincer, Scott Lundberg, Matt Kaeberlein, Su-In Lee

## Abstract

Prior studies on all-cause mortality traditionally use linear models; however, growing field of explainable artificial intelligence (XAI) can improve prediction accuracy over traditional linear models using complex machine learning (ML) models while still revealing novel insights. We propose the IMPACT (Interpretable Machine learning Prediction of All-Cause morTality) framework that implements and explains complex, non-linear ML models by combining a tree ensemble mortality prediction model and a principled XAI technique. We apply IMPACT to the NHANES (1999-2014) dataset, which enables us to understand different subpopulations according to shorter or longer term mortality and younger and older individuals. Our IMPACT models have higher predictive accuracy than popular pre-existing mortality risk scores and biological ages. Using individualized feature importance scores, we discover novel risk predictors (e.g., arm circumference) and interactions between risk predictors (e.g., serum chloride with age and/or gender). Furthermore, IMPACT provides a novel perspective of reference intervals and may suggest that the widely accepted reference intervals for serum albumin, mean cell volume and platelet count may in fact be sub-optimal for health. Finally, in order to ensure that our models are useful to as broad of a community as possible, we develop and publish a variety of explainable risk scores usable by individuals with and without medical expertise. The predictive accuracy of IMPACT combined with the capability of discovering mortality risk predictors and complex relationships demonstrates the value and utility of XAI in epidemiologic study design.

## 1 Main

Identification of risk factors and prediction of all-cause mortality have long been central issues in epidemiology. Most prior studies identify risk factors using associations between each predictor and mortality [3, 23, 31]; only a few papers used multi-variate linear models to predict mortality and identify risk factors [55, 12]. In terms of prediction, a variety of linear mortality risk scores have been proposed to help differentiate unhealthy individuals [18, 11, 48]. Although linear models have historically been popular because they are interpretable, modern complex machine learning (ML) models often achieve higher predictive accuracy because they capture interactions among variables in addition to non-linear relationships, such as “U-shaped” relationship.

The field of artificial intelligence (AI) has seen significant advances in *supervised learning* problems, which involve predicting an *outcome variable* (e.g., all-cause mortality) based on a set of *features* (e.g., individual-level characteristics). Notable applications of AI in healthcare include diabetic retinopathy detection in ophthalmology images [15], lung cancer classification from histopathology images [6] and skin cancer classification [8]. Despite this progress, a major obstacle to the adoption of AI in healthcare is that many of them are considered “black box”, which refers to the lack of *interpretability*. The inability to understand why a model makes a prediction is especially harmful in healthcare applications where the patterns a model discovers can be even more important than its predictive accuracy. This is especially true in epidemiology, which aims to identify important variables to guide public health policy or detect risk predictors that warrant further study. To address this need, we turn to a variety of techniques to help understand complex ML models from the emerging area of explainable AI (XAI) [44, 27, 29].

We combine an accurate, complex ML model and a state-of-the-art XAI technique to predict and understand all-cause mortality. To our knowledge, our study is the first to use complex ML models to do a systematic and integrated study of the associations between a large number of variables and all-cause mortality. We present the IMPACT (Interpretable <u>M</u>achine learning <u>P</u>rediction of <u>A</u>ll-<u>C</u>ause mor<u>T</u>ality) framework (Figure 1) and apply it to the NHANES (1999-2014) dataset to reveal novel all-cause mortality findings. First, using explainable complex ML models rather than linear models, we identify new risk predictors that are highly informative of future mortality. Second, our flexible models capture non-linear relationships which provide more comprehensive information about the relationship between feature values and mortality risk: for example, the “inflection” points of risk predictors could provide a novel perspective of reference intervals and have significant implications in public health. Third, interpretability points us to the most important features which enable us to develop highly accurate, efficient (using less features) and interpretable mortality risk scores. Furthermore, the individualized explanation of risk scores can help users understand their biggest risk factors and adjust their lifestyle. Finally, IMPACT risk scores (Supplementary Table 2) have higher predictive power than popular mortality risk scores [18, 11, 12, 48] and biological ages [19, 16, 24, 26] (Table 1). All our results and risk scores are available in an interactive website^1^ in order to encourage exploration of important risk predictors and to support the use of interpretable individual risk scores for both individuals with and without medical expertise.

**Figure 1:**
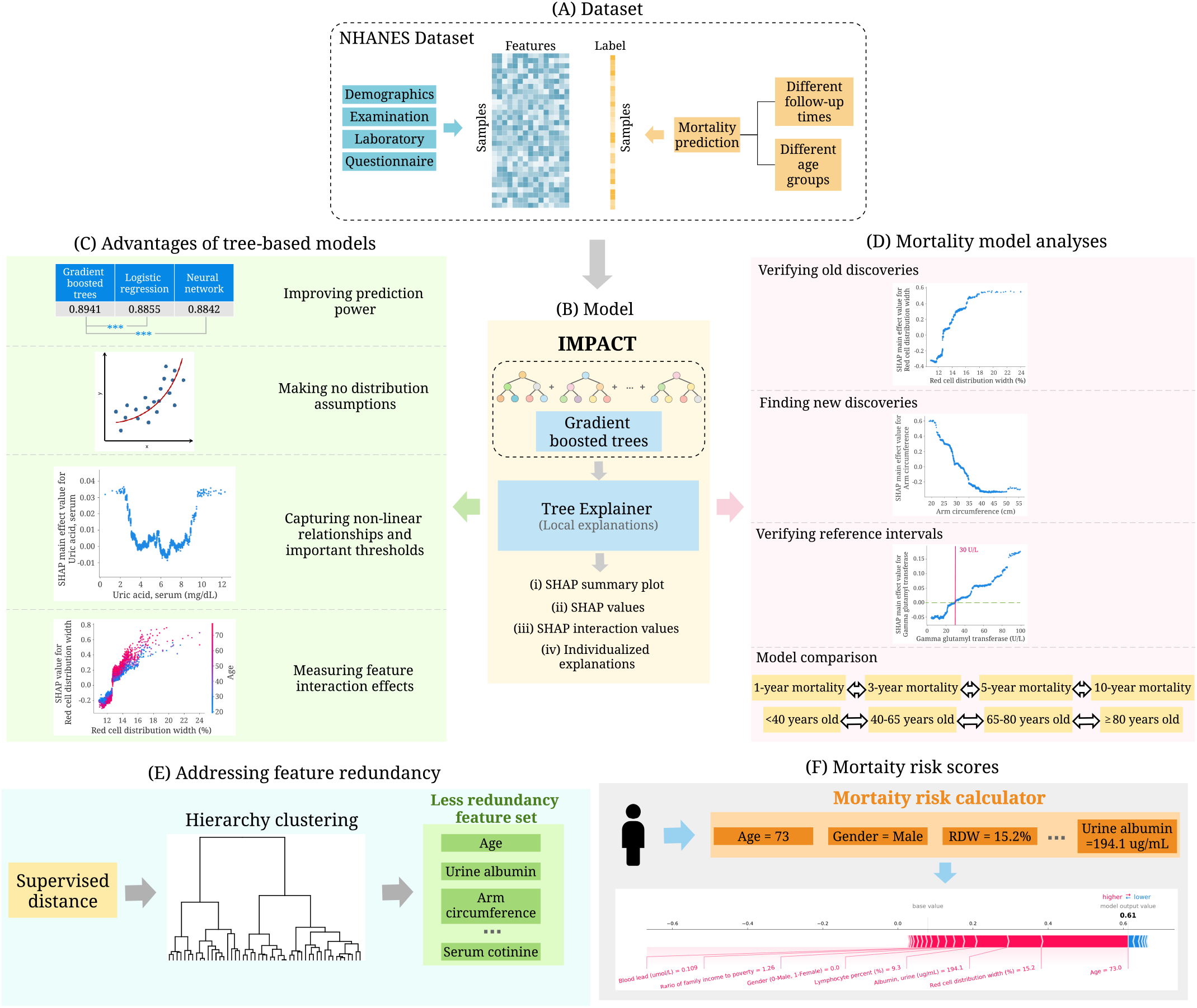
Overview of the IMPACT model and analyses. (A) We use the NHANES (1999-2014) dataset which includes 151 variables and 47,261 samples. The variables can be categorized into four groups: demographics, examination, laboratory and questionnaire. We train the model using different follow-up times and different age groups. (B) IMPACT combines tree-based models with an explainable AI method. Specifically, IMPACT (1) trains tree-based models for mortality prediction using NHANES dataset (2) uses TreeExplainer to provide local explanations for our models. (C) We illustrate the advantages of interpretable tree-based models compared to traditional linear models in epidemiological studies. (D) We further analyze all mortality models and demonstrate the effectiveness of IMPACT to verify existing findings, identify new discoveries, verify reference intervals, obtain individualized explanations, and compare models using differ-ent follow-up times and age groups. (E) We propose a supervised distance which helps us explore feature redundancy. We develop a supervised distances-based feature selection method which helps us select predictive and less-redundant features. (F) We build mortality risk scores that are applicable to professional and non-professional individuals with different cost-vs-accuracy tradeoffs. The individualized explanations of IMPACT shows the impact of each risk factor for the risk score.

**Table 1:** Comparing the predictive power of popular mortality risk scores and biological ages with IMPACT. The “AUC” column shows the AUCs reported in the original paper. The “AUC of IMPACT” column shows the AUCs of the IMPACT model trained with all features. The “AUC of IMPACT-20” column shows the IMPACT model trained with the selected top 20 features (Method 6). The “HR” column shows the hazard ratios for all-cause mortality with a standard unit (z-score) increase in epigenetic age acceleration reported in the original paper. The “HR of IMPACT” column shows the hazard ratio for all-cause mortality with a standard unit increase in the IMPACT score trained with all features. The “HR of IMPACT-20” column shows the hazard ratio for all-cause mortality with a standard unit increase in the IMPACT score trained with the selected top 20 features. (**) represents a P-value < 0.01.

|  | Task | Age | AUC | AUC of IMPACT | AUC of IMPACT-20 |
| --- | --- | --- | --- | --- | --- |
| Mortality risk scores |  |  |  |  |  |
| Intermountain [18] | 1-year mortality | 18+ | 0.84 | 0.92 | 0.92 |
| Gagne Index [11] | 1-year mortality | 65+ | 0.79 | 0.84 | 0.86 |
| Intermountain [18] | 5-year mortality | 18+ | 0.87 | 0.89 | 0.89 |
| Prognostic score [12] | 5-year mortality | 40-70 | Male: 0.80 | Male: 0.83 | Male: 0.82 |
|  |  |  | Female: 0.79 | Female: 0.81 | Female: 0.80 |
| Schonberg Index [48] | 5-year mortality | 65+ | 0.75 | 0.81 | 0.80 |
| Biological ages |  |  |  |  |  |
| Horvath DNAm Age [19, 24] | 10-year mortality | 21-84 | 0.56 | 0.92 | 0.91 |
| Hannum DNAm Age [16, 24] | 10-year mortality | 21-84 | 0.57 | 0.92 | 0.91 |
| DNAm PhenoAge [24] | 10-year mortality | 21-84 | 0.62 | 0.92 | 0.91 |
| Phenotypic Age [24, 25] | 10-year mortality | 20-85 | 0.88 | 0.92 | 0.91 |
|  | Task | Age | HR | HR of IMPACT | HR of IMPACT-20 |
| Horvath DNAm Age [19, 33] | Cox regression (mortality) | 50+ | 1.03 | 2.40 ** | 2.35 ** |
| Hannum DNAm Age [16, 33] | Cox regression (mortality) | 50+ | 0.92 | 2.40 ** | 2.35 ** |
| DNAm PhenoAge [24, 33] | Cox regression (mortality) | 50+ | 1.13 | 2.40 ** | 2.35 ** |
| GrimAge [26, 33] | Cox regression (mortality) | 50+ | 1.91 ** | 2.40 ** | 2.35 ** |

## 2 Results

### 2.1 Data cohorts

This study includes the NHANES ^1^ data samples between 1999-2014. We include demographic, laboratory, examination, and questionnaire features that could be automatically matched across different NHANES cycles. After data preprocessing (Method 1), 47,261 samples with 151 features (Supplementary appendix 2) remain. Follow-up mortality data is provided from the date of survey participation through December 31, 2015. We predict all-cause mortality for two broad categories: (1) follow-up times of 1-year, 3-year, 5-year, and 10-year and (2) age groups of <40, 40-65, 65-80, and 80 years old. For different age groups, we predict 5-year mortality. The dataset is randomly divided into training (80%) and testing (20%) sets. The demographic characteristics and sample size of the data for different tasks are shown in Figure 2.

**Figure 2:**
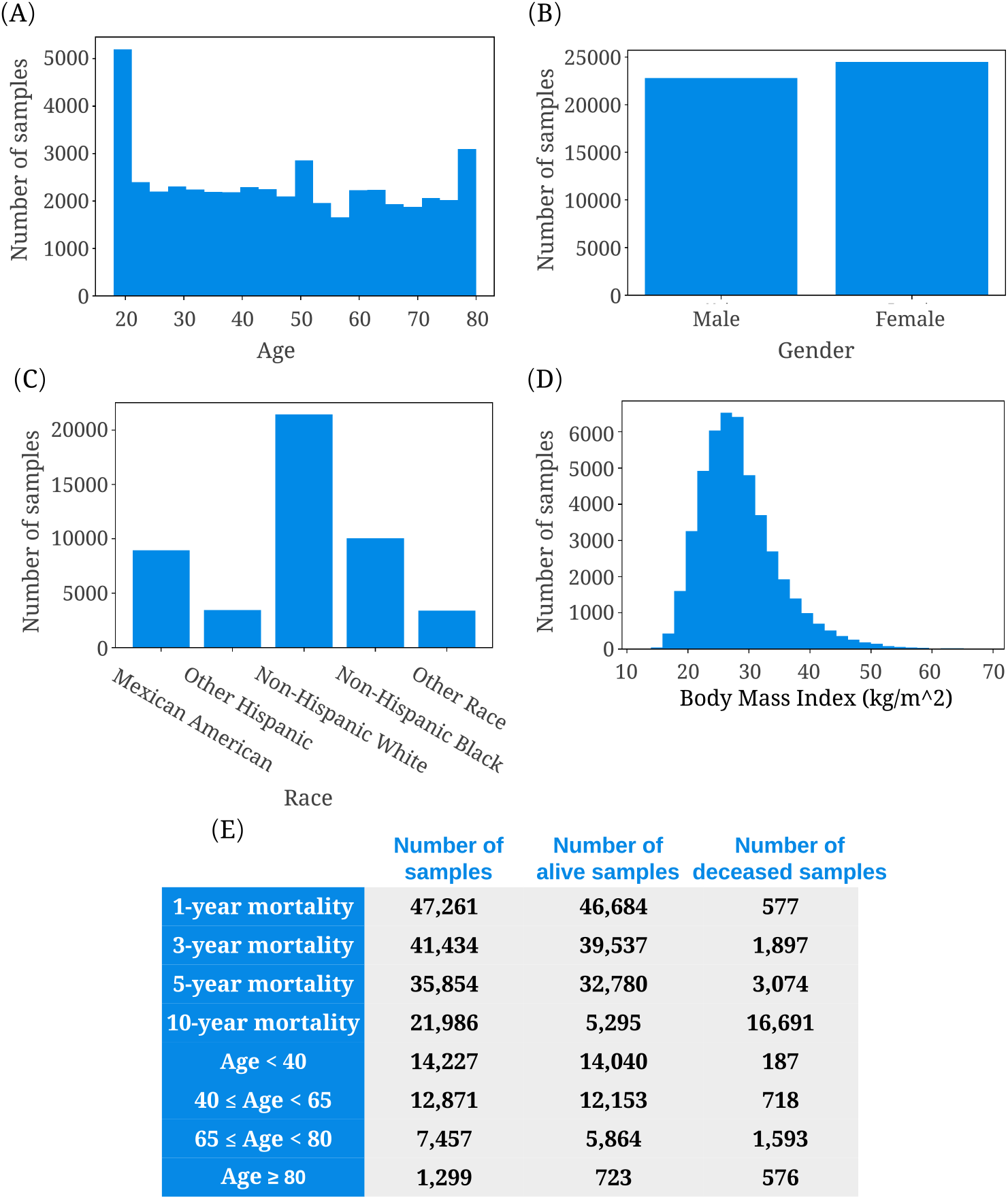
(A)-(D) Histograms of age, gender, race, and body mass index in the NHANES dataset. (E) The sample size and number of living and deceased samples for different follow-up times and different age groups. For different age groups, the follow-up time is set to 5 years.

### 2.2 IMPACT framework

To achieve the high-accuracy and explainable mortality prediction models, we present IMPACT (Figure 1) framework, which combines tree-based models and TreeExplainer [28]. To model all-cause mortality, we use gradient boosted trees (GBTs). GBTs are nonparametric methods composed of iteratively trained decision trees. The final ensemble of trees captures non-linearity and interactions between predictors. The hyperparameters are chosen by GridSearch and 5-fold cross-validation (Method 2). The performance of the models is measured with the area under the receiver operator characteristic curve (AUROC).

To explain the GBT models, we utilize TreeExplainer [28], which provides a local (i.e., for each subject) explanation of the impact of input features on individual predictions (Method 3). Specifically, TreeExplainer calculates exact SHAP [27] (SHapley Additive exPlanations) values, which guarantee a set of desirable theoretical properties. First, SHAP values are *additive*. They sum to the model’s output, i.e., the log-odds for GBTs. Second, they are *consistent*, which means features that are unambiguously more important are guaranteed to have a higher SHAP value. Therefore, SHAP values are consistent and accurate calculations of each feature’s contribution to the model’s prediction. In our study, higher SHAP values imply large contributions to mortality risk. TreeExplainer also extends local explanations to capture feature interactions directly. By showing the impact of each variable and interactions between variables for local, sample-specific explanations, we can obtain a comprehensive understanding of why the model made a specific mortality prediction.

### 2.3 Advantages of tree-based models

Linear models are commonly used in epidemiology studies because their coefficients indicate each feature’s contribution to the model’s prediction [35]. However, more expressive models, such as tree-based models, can achieve higher predictive accuracy for many datasets by learning non-linear relationships between features and the outcome variable. Gradient boosted trees (GBTs) have achieved state-of-the-art performance in many domains [9, 51, 43, 62]. We observe the same trend in our study: tree-based models outperform both linear models and neural networks across all tasks we consider (Figure 3A). The superior prediction performance of tree models indicates that we can capture signals relevant to mortality, which alternative approaches could not. Besides predictive power, tree-based models have more advantages compared with traditional linear models. Our study illustrates the advantages of tree-based models in epidemiology, including making minimal assumptions, capturing non-linear relationships, important thresholds and interaction effects.

**Figure 3:**
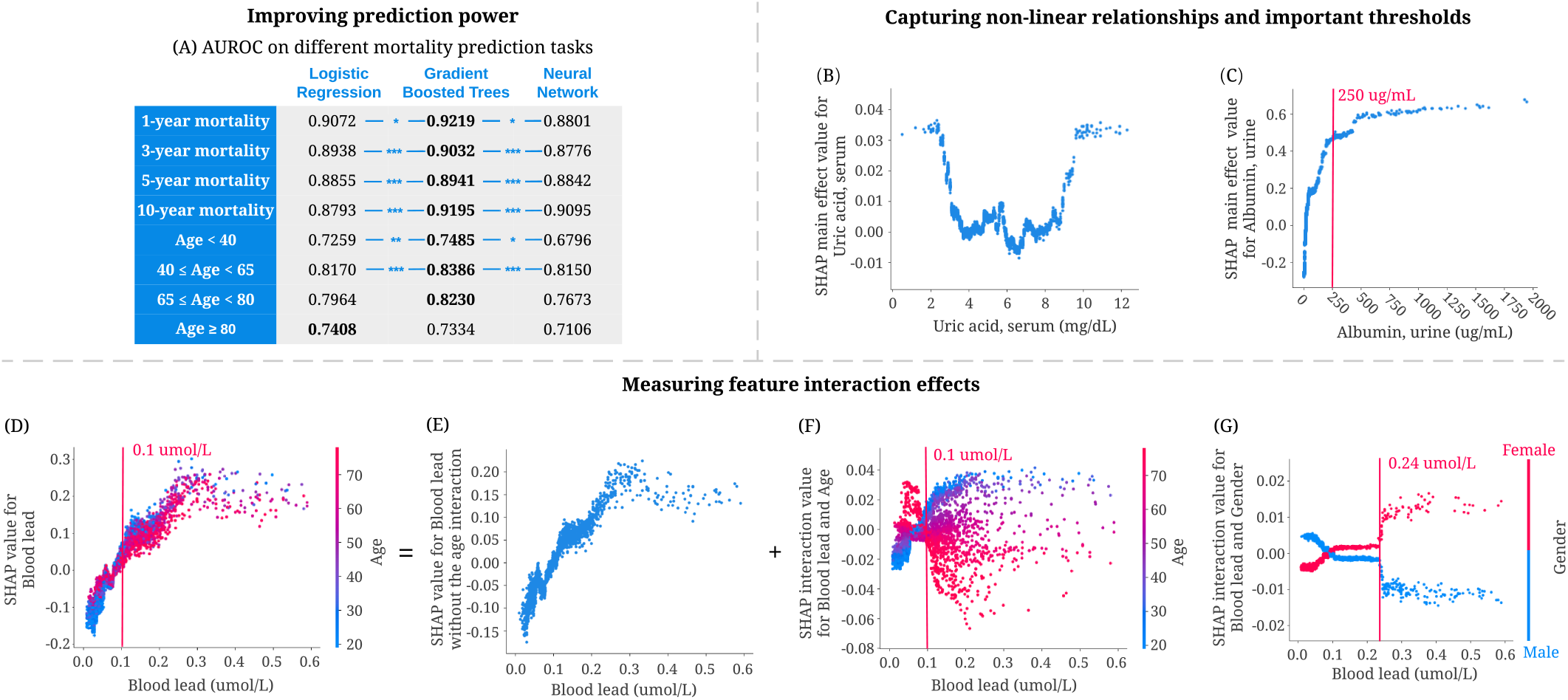
Advantages of tree-based models for mortality prediction. (A) The area under the ROC curve (AUROC) of gradient boosted tree models outperforms both linear models and neural networks for six of our prediction models.(* * *) represents a P-value < 0.001, (**) represents a P-value < 0.01, and (*) represents a P-value < 0.05. P values are computed using bootstrap resampling over the tested time points while measuring the difference in area between the curves. (B,C) Tree-based models can capture non-linear relationships and important thresholds. (B) The main effect of uric acid on 5-year mortality. Higher SHAP value leads to higher mortality risk (C) The main effect of urine albumin on 5-year mortality. (D–G) Tree-based models can measure feature interaction effects. (D) SHAP value for blood lead level in the 5-year mortality model. Each dot corresponds to an individual. The color corresponds to the value of a second feature (i.e. age) that has an interaction effect with blood lead. (E) We can use SHAP interaction values to remove the interaction effect of age from the model and obtain the SHAP value of blood lead without the age interaction on 5-year mortality. (F) Plotting just the interaction effect of blood lead with age shows how the effect of blood lead on mortality risk varies with age. (G) The SHAP interaction value of blood lead vs. gender in the 5-year mortality model.

#### Tree-based models make minimal assumptions about the data distribution

Several assumptions associated with linear models (e.g., linearity, independence, normality, etc.) restrict the features linear models can use. To satisfy these assumptions, scientists often manually transform non-linear variables before fitting a model (e.g., log-transformation, discretization of continuous variables, etc.). For instance, to explore the effect of blood lead on mortality, researchers first discretized blood lead using different thresholds. They observed that individuals with blood lead levels higher than the threshold had increased mortality risk compared to those with lower blood lead levels [30, 34, 47]. In comparison, tree-based models make minimal assumptions about the data distribution and need no data transformations. Figure 3D shows a positive relationship between blood lead and 5-year mortality risk. Tree-based models can capture complex relationships directly without the need of manually transforming the variables.

#### Tree-based models capture non-linear relationships and important thresholds

Discovering non-linear relationships is important but challenging for epidemiological research using traditional linear models. J-shaped and U-shaped associations are two common and meaningful non-linear relationships [32]. However, linear models must use manually transformed features to capture non-linear relationships. As an example, Suliman et al. showed a J-shaped relationship between uric acid levels and mortality in patients with stage 5 chronic kidney disease (CKD) using a linear model by dividing uric acid level into three categories and calculating the hazard ratio for each. Unlike linear models, tree-based approaches can directly capture non-linear relationships. We observe a U-shaped relationship between uric acid level and all-cause 5-year mortality predictions in Figure 3B. This relationship differs from the J-shaped one in previous work, possibly because of categorization, which loses essential information about values within the categories.

Additionally, discovering thresholds (i.e., inflection points beyond which changing a feature’s value has diminishing returns) is significant in epidemiological analysis. Figure 3C shows that 250 *μ*g/mL is an important threshold: according to our model, increasing urine albumin generally increases 5-year mortality risk; however, urine albumin higher than this threshold has almost the same impact on mortality risk.

#### Tree-based models measure feature interaction effects

Feature interaction examines how the effect of one feature on the outcome differs across strata of another feature and shows the complex relationship of two features on the outcome [7]. Tree-based models can naturally capture interaction effects by splitting on different features in the same tree. As shown in Figure 3D-F, SHAP dependence plots can be decomposed into main effects and interaction effects for each sample. Figure 3F highlights a specific interaction: the relationship of blood lead level to mortality presents differently for young and old individuals. Specifically, for those with blood lead higher than 0.1 *μ*mol/L, younger individuals have a higher 5-year mortality risk than older individuals. Figure 3G shows the SHAP interaction effects of gender with blood lead level: females have a higher 5-year mortality risk than males with blood lead levels higher than 0.24 *μ*mol/L. The interaction effects of age and gender with blood lead level cannot be clearly identified without SHAP interaction values because being male or older generally increases mortality risk. These findings highlight how the interaction effects detected by our model open opportunities for further research.

### 2.4 Discoveries from 5-year mortality prediction

Figure 4A shows a summary plot that displays the magnitude, prevalence, and direction of the effect of the top 20 most impactful features on 5-year mortality prediction (Method 4). This summary plot provides an integrated explanation of the 5-year IMPACT model. Several features have previously been shown to be associated with mortality in epidemiological studies. Our results examine and support these studies’ conclusions as well as surface additional discoveries, including novel features, thresholds, and non-linear relationships.

**Figure 4:**
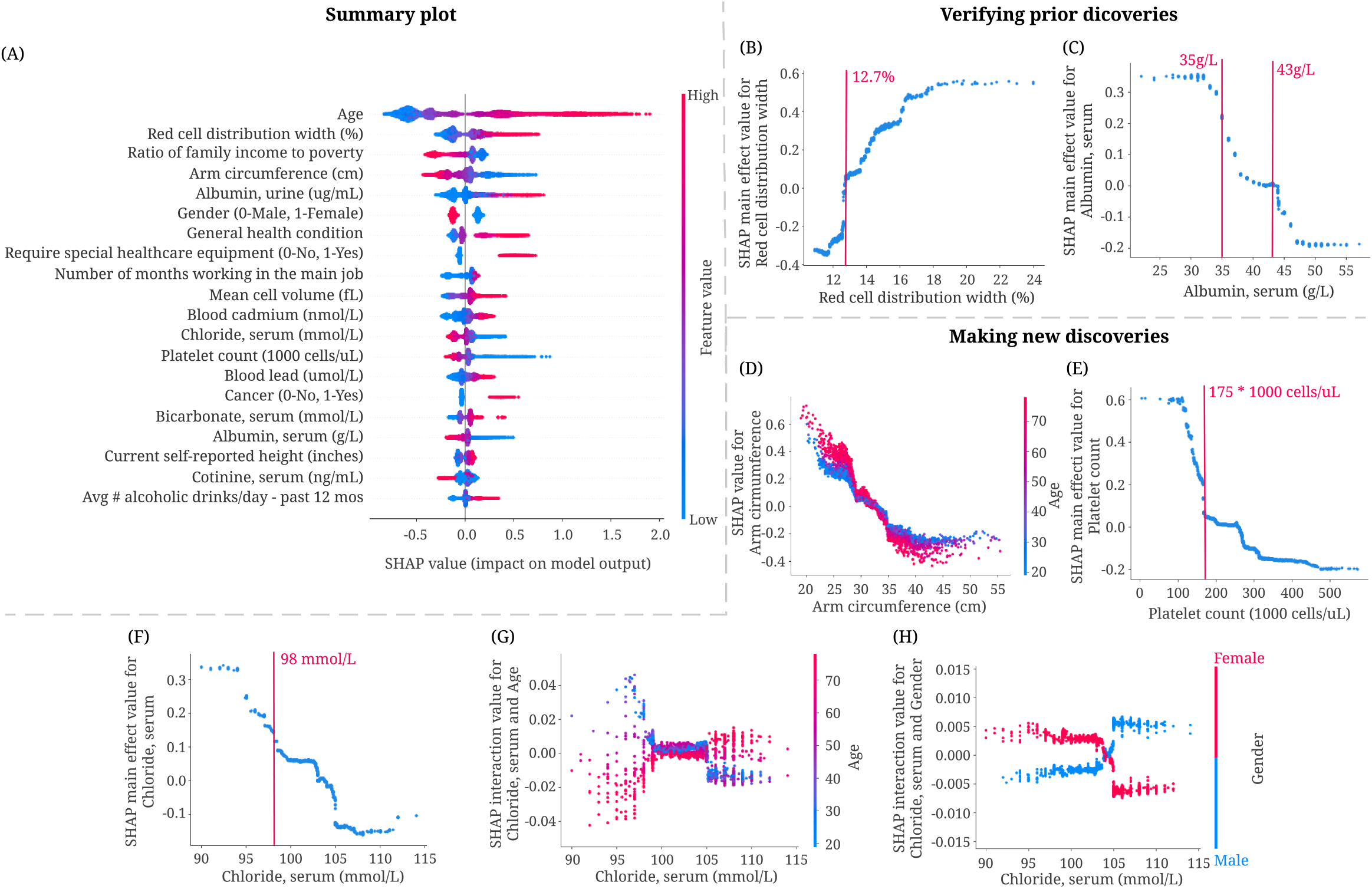
Combining 5-year mortality prediction gradient boosted trees models and local explanations to achieve significant discoveries about the entire model and individual features. (A) SHAP summary plot for the gradient boosted trees trained on the 5-year mortality prediction task. The plot shows the most impactful features on prediction (ranked from most to least important) and the distribution of the impacts of each feature on the model output, which includes a set of plots where each dot corresponds to an individual. The colors represent feature values for numeric features: red for larger values, and blue for smaller. The thickness of the line that is comprised of individual dots is determined by the number of examples at a given value. A negative SHAP value (extending to the left) indicates reduced mortality risk, while a positive one (extending to the right) indicates increased mortality risk. (B,C) IMPACT can verify well-studied features associated with mortality. (B) The main effect of red cell distribution width on 5-year mortality. (C) The main effect of serum albumin on 5-year mortality. (D-H) IMPACT can identify less well-studied features associated with mortality. (D) The SHAP value for arm circumference in 5-year mortality model. (E) The main effect of platelet count on 5-year mortality. (F) The main effect of serum chloride on 5-year mortality. (G) The SHAP interaction value of serum chloride vs. age in the 5-year mortality model. (H) The SHAP interaction value of serum chloride vs. gender in the 5-year mortality model.

#### IMPACT verifies well-studied features associated with mortality

Some of the top 20 most important features for our 5-year mortality prediction models have been previously identified. For example, red cell distribution width (RDW), the second most important feature of the 5-year IMPACT model, has been shown to have a strong positive relationship with mortality by many studies under several conditions [10, 39, 40, 41]. We also observe the positive relationship between RDW and risk of mortality (Figure 4B); moreover, 12.7% is an important threshold over which RDW manifests a positive effect on mortality. Serum albumin level’s relation to mortality is also well-studied. Previous studies show that serum albumin is negatively associated with mortality risk [5, 13, 42]. The relationship shown in Figure 4C matches this trend. Furthermore, Corti et al. showed that serum albumin level<35 g/L was associated with a significantly increased risk of mortality compared to serum albumin levels greater than 43 g/L [5]. We observe that 35 g/L and 43 g/L are indeed key inflection points (Figure 4C): serum albumin levels lower than 43 g/L have a positive relationship with mortality prediction, while those around 35 g/L are associated with a dramatically increased mortality risk.

#### IMPACT identifies less well-studied features associated with mortality

Some of the top 20 most important features identified by IMPACT are less appreciated as mortality risk factors in the existing epidemiological literature. Three of these are arm circumference, platelet count, and serum chloride level. Figure 4D shows a negative relationship between arm circumference and 5-year mortality, especially for older people. This negative relationship is consistent with previous work [2, 64]. IMPACT ranks arm circumference as the fourth most important feature for 5-year mortality prediction, with an importance ranking that significantly exceeds that of BMI (the 56th). This suggests that smaller arm circumference is more predictive than BMI for modeling mortality, as in [52].

Figure 4E shows a negative relationship between platelet count, the 13th most important feature, and 5-year mortality. 175 × 1, 000 cells/*μ*L is an important threshold; platelet count lower than that level is associated with dramatically increased mortality risk. Serum chloride is also inversely related to 5-year mortality (Figure 4F). The normal adult value for chloride is 98-106 mmol/L. We observe that serum chloride lower than 98 mmol/L is associated with sharply increased mortality risk. In Figure 4G–H, we plot the interaction effect of age and sex with serum chloride level. This analysis reveals that younger people and females with low serum chloride have a higher mortality risk than older people and males. The interaction effect of age and serum chloride shows that early rather than late-onset low chloride level has a greater effect on the model.

#### IMPACT can purovide additional perspective to laboratory reference intervals

Reference interval (RI) is the range of values that is deemed normal for a physiologic measurement in healthy persons [22] It is the most common decision support tool to interpret patient laboratory test results. RIs enable differentiation of healthy and unhealthy individuals [38, 21]. Hence, the quality of the RIs is as crucial as the quality of the result itself. RIs in use today are most commonly defined as the central 95% of laboratory test results in a reference population. Unfortunately, this definition does not consider mortality or disease risk, which may lead to misdiagnosis since RIs are often used to identify unhealthy individuals. The partial dependence plots of IMPACT models directly reflect the effects of the features on mortality risk, which provides an alternative perspective for identifying inappropriate reference intervals with mortality/disease relevance.

We define the relative risk percentage (RRP) that measures the relative risk (Method 1 3.3) of the feature values within the reference interval compared to the relative risk of all values (Table 2). A higher RRP indicates that the feature values within the reference interval may lead to high mortality risk, which we need to pay special attention to. The first four features in Table 2 have relatively low 5-year mortality RRP. From Figure 5A-D, we observe that the values of these features within the reference interval have low 5-year relative mortality risk; the values outside reference interval may lead to increased 5-year mortality risk. Therefore, IMPACT confirms the reference intervals of these four features as optimal for mortality risk. In contrast, the RRP of the last four features in Table 2 are high. Figure 5E-H also shows that the relative 5-year mortality risk of the values within the reference interval is high compared to the maximum relative risk of all values. Hence, IMPACT identified the divergence where reference intervals appear to be poorly tuned to mortality risk, suggesting that these reference intervals may in fact be sub-optimal for health.

**Table 2:**
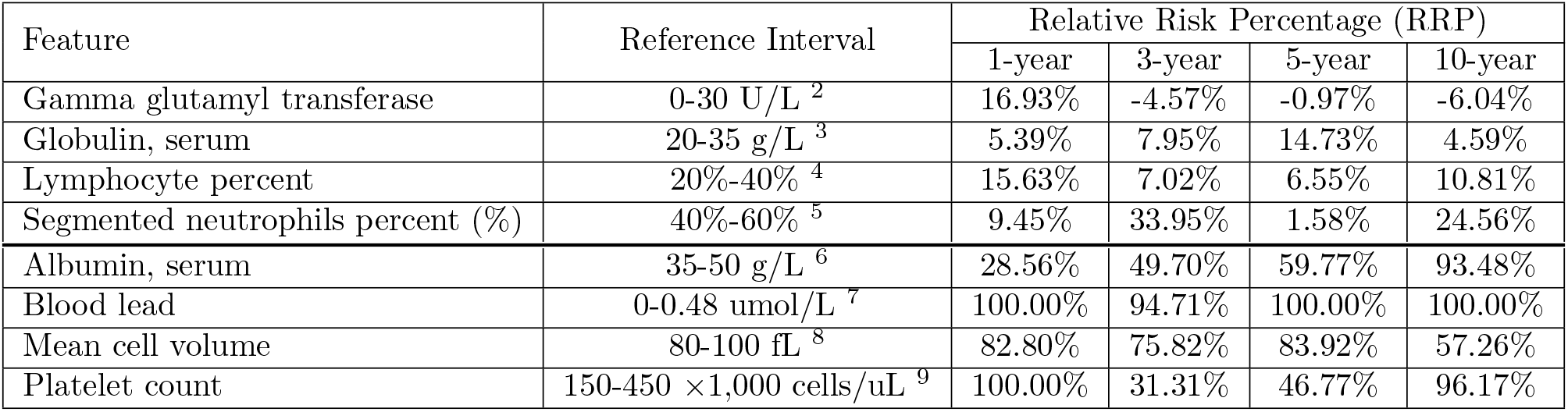
Providing additional perspective to laboratory reference intervals. The table lists the reference interval and relative risk percentage (RRP; Method 1 3.3) of the selected laboratory features. RRP measures the relative risk of the feature values within the reference interval compared to the relative risk of all values.

| Feature | Reference Interval | Relative Risk Percentage (RRP) |  |  |  |
| --- | --- | --- | --- | --- | --- |
|  |  | 1-year | 3-year | 5-year | 10-year |
| Gamma glutamyl transferase | 0-30 U/L <sup>2</sup> | 16.93% | -4.57% | -0.97% | -6.04% |
| Globulin, serum | 20-35 g/L <sup>3</sup> | 5.39% | 7.95% | 14.73% | 4.59% |
| Lymphocyte percent | 20%-40% <sup>4</sup> | 15.63% | 7.02% | 6.55% | 10.81% |
| Segmented neutrophils percent (%) | 40%-60% <sup>5</sup> | 9.45% | 33.95% | 1.58% | 24.56% |
| Albumin, serum | 35-50 g/L <sup>6</sup> | 28.56% | 49.70% | 59.77% | 93.48% |
| Blood lead | 0-0.48 umol/L <sup>7</sup> | 100.00% | 94.71% | 100.00% | 100.00% |
| Mean cell volume | 80-100 fL <sup>8</sup> | 82.80% | 75.82% | 83.92% | 57.26% |
| Platelet count | 150-450 ×1,000 cells/uL <sup>9</sup> | 100.00% | 31.31% | 46.77% | 96.17% |

**Figure 5:**
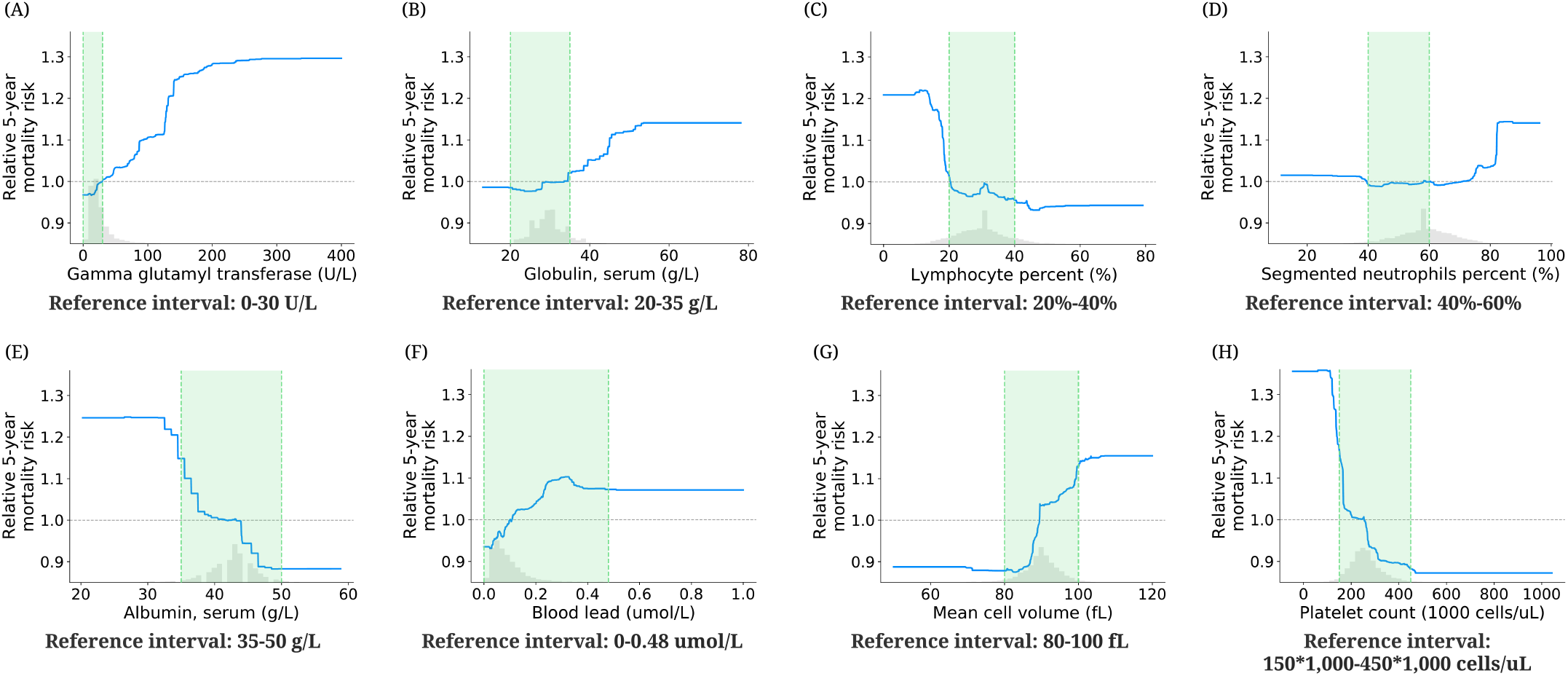
Effect of varying laboratory feature values on 5-year mortality risk. These partial dependence plots show the change in relative 5-year mortality risk (Method 1 3.3) for all values of a given laboratory feature. The grey histograms on each plot show the distribution of values for that feature in the test set. The green shaded region shows the reference interval of each feature.

### 2.5 Discoveries for mortality prediction using different follow-up times

The relationship between each feature and mortality may change for different models. For instance, comparing important features between IMPACT models using different follow-up times can reveal features that are only predictive of short-term mortality, not longer-term mortality (and vice versa).

#### IMPACT identifies trends for 1-year, 3-year, 5-year and 10-year mortality prediction models

Figure 6A shows the top 20 most important features and relative importance of input features in IMPACT’s 1-year, 3-year, 5-year, and 10-year mortality prediction models. Feature importance rankings change significantly between these four models. Some features are important for all four (e.g., age, RDW, and urine albumin level). Some features become more important over time (e.g., platelet count, whose importance ranking is 75 for the 1-year model and 12 for the 10-year model). Other features become less important over time (e.g., serum potassium, whose importance ranking is 17 for the 1-year model and 42 for the 10-year model). These results provide a more comprehensive understanding of shorter- and longer-term mortality risk, which can facilitate the investigation of mechanisms underlying risk predictors and potentially help validate interventions.

**Figure 6:**
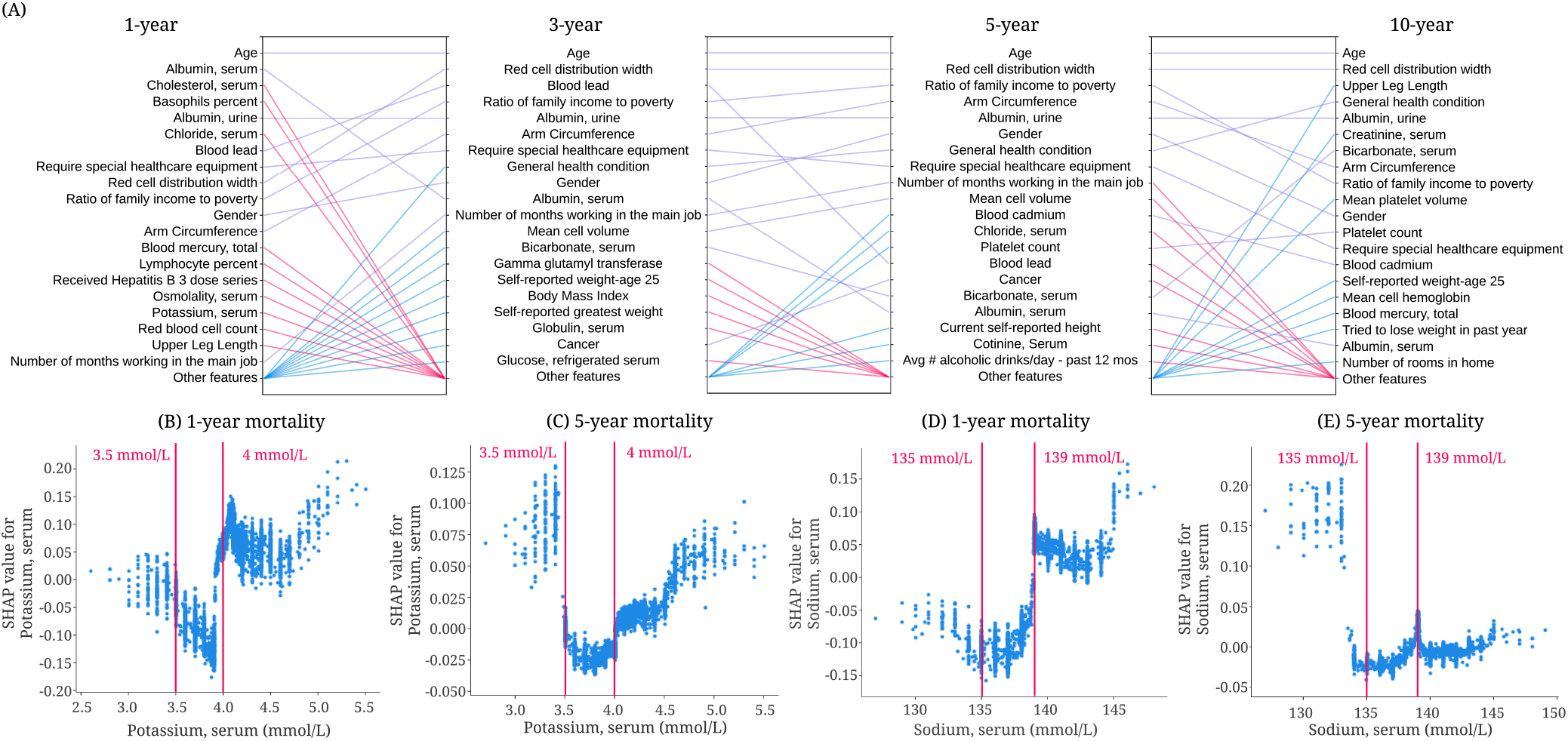
Identifying important discoveries for mortality prediction from tree-based models using different follow-up times. (A) Relative importance of input features in 1-, 3-, 5- and 10-year mortality models. For each model, the figure shows the 20 most important features of prediction (ordered by the importance). The purple line indicates that the feature is in the top 20 features of two models. Blue and red lines indicate the feature is in the top 20 features of one model, but not in the top 20 features of the other. (B) The SHAP value of serum potassium in the 1-year mortality model. (C) The SHAP value of serum potassium in the 5-year mortality model. (D) The SHAP value of serum sodium in the 1-year mortality model. (D) The SHAP value of serum sodium in the 5-year mortality model.

The relationship between each feature and mortality may change for models that predict different mortality outcomes or utilize different subsamples of the general population. For instance, Figure 6B-C shows the SHAP value for serum potassium in IMPACT’s 1-year and 5-year mortality prediction models. The finding that serum potassium lower than 3.5 mmol/L and higher than 4.0 mmol/L are associated with increased mortality risk has been previously observed [1, 14, 36]. Interestingly, for the 1-year model, hyperkalemia (high potassium) has a higher mortality risk than hypokalemia (low potassium). For the 5-year model, hypokalemia has the same or higher mortality risk than hyperkalemia. Figure 6D shows that serum sodium higher than 139 mmol/L increases 1-year mortality risk, and low serum sodium with negative SHAP values decreased mortality risk. However, the relationship differs completely in the 5-year mortality prediction model (Figure 6E): hyponatremia (serum sodium <135 mmol/L) is associated with a higher 5-year mortality risk. This type of insight, especially regarding the differences of non-linear trends, is not apparent using linear models.

Likewise, we can compare models trained on distinct subpopulations (e.g., samples in different age groups). The differences between these models can help researchers identify risk predictors relevant to each subpopulation. Comparing models in this way can provide epidemiological insights that may guide policy for specific at-risk populations. The discoveries for mortality prediction using different age groups are discussed in Supplementary appendix 1.

### 2.6 Exploring feature redundancy using supervised distance

Often features in datasets are partially or fully redundant with each other, in the sense that a model could use either feature and still achieve the same accuracy. It is important to be aware of redundant features when we interpret a model because these features may include the same information about the output and thereby split the importance of this information. To this end, we propose a supervised distance, which helps us explore and better understand redundant features (Method 5). Building upon supervised distance, we develop a feature selection method to maximize accuracy and minimize redundancy.

#### Supervised distances measures feature redundancy and identifies redundant groups of features

Researchers often use unsupervised methods such as some form of correlation-based clustering to find dependent features [61, 54]. However, when we have a specific prediction task in mind, we would like to measure the feature redundancy with respect to the task. The supervised distance can be an accurate measure of this feature redundancy. (Method 1 5.1). Specifically, supervised distance measures the similarity of the two features’ information about the prediction task. It is scaled roughly between 0 and 1, where 0 distance means the features are perfectly redundant regarding the prediction task and 1 means they are not redundant at all.

To identify groups of redundant features, we hierarchically cluster all features according to supervised distance (Supplementary Figure 3; Method 1 5.1). Redundant features that have the same information about the output group together. For example, arm circumference, the fourth most important feature of 5-year IMPACT model, is grouped with weight-related features: BMI, waist circumference, weight, etc. These weight-related features all contain similar information about 5-year mortality. To further explore the predictive ability of the features, we train models using one weight-related feature and all non-weight-related features (**reducing redundancy models**) and models using one weight-related feature in addition to age and gender (**single feature models**) (Method 1 5.2). Arm circumference is the most predictive weight-related feature across all settings (Figure 7A). These results indicate that arm circumference may be more informative than other weight-related features with respect to all-cause mortality. Another example would be the cluster that includes many blood test features (Figure 7B). Similar to arm circumference, serum albumin is the most predictive feature among these blood test features. In summary, using supervised distance, we can easily identify redundant feature groups and select the most representative feature based on predictive power. These selected features can be the strongest risk predictors because they have strong predictive power and can represent a number of features.

**Figure 7:**
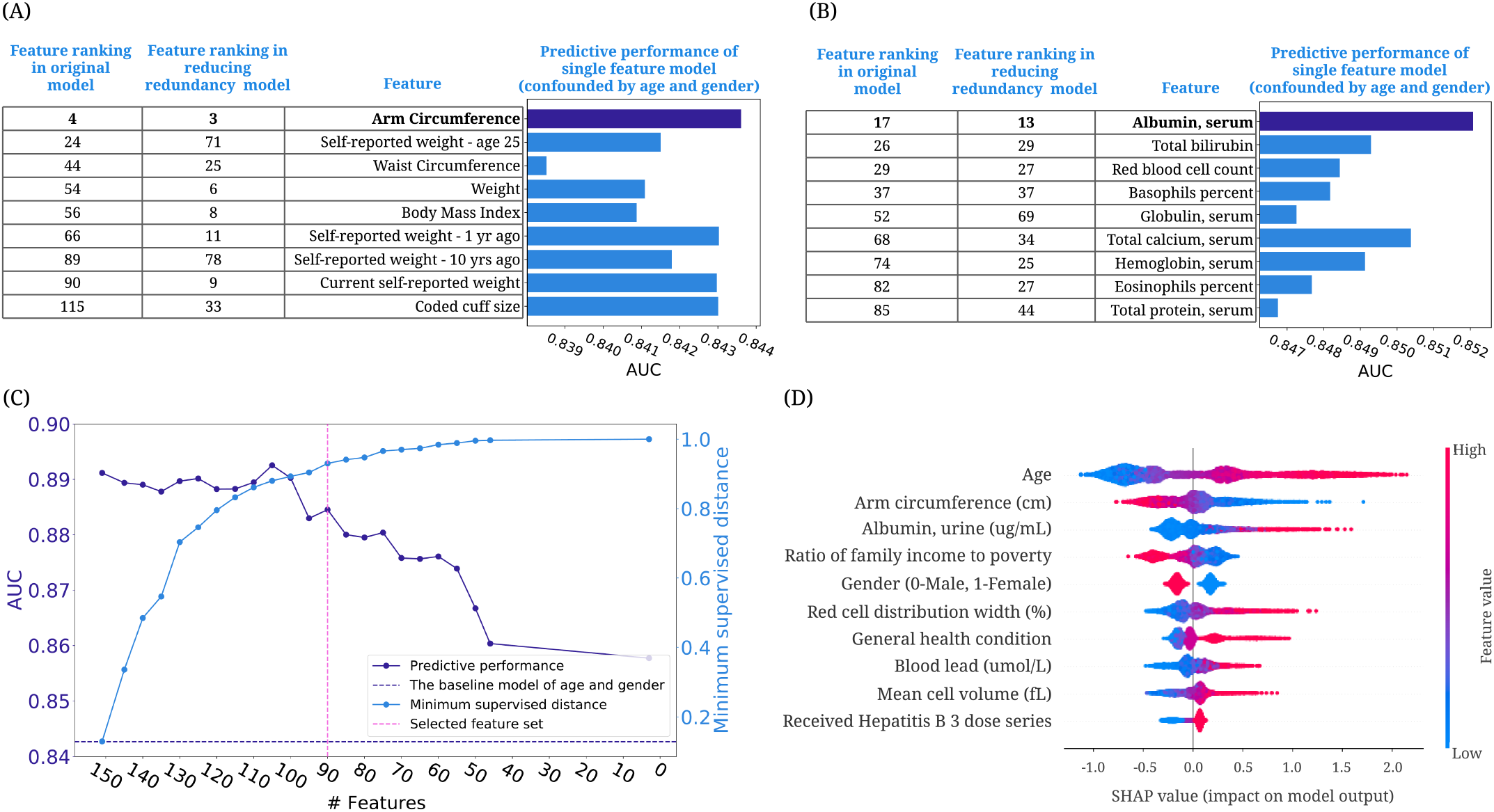
Exploring features redundancy using supervised distance. (A) The feature importance ranking of the BMI-related features in original models and reducing redundancy models, and the AUC of the single feature models confounded by age and gender. (Method 1 5.2) (B) The feature importance ranking of the selected laboratory features in original models and reducing redundancy models and the AUC of the single feature models confounded by age and gender. (C) The AUC of the models using the selected feature sets and the minimum feature redundancy within the selected feature sets when running supervised distance-based feature selection. The purple dashed line shows the AUC of the model trained on age and gender. The pink dashed line indicates the feature set we select for further analysis. (D) SHAP summary plot for the gradient boosted trees trained on the selected 90 features for the 5-year mortality prediction.

#### Supervised distance-based less-redundant feature selection

To better address the redundancy in the dataset, we propose a recursive feature selection method based on the supervised distance to select the predictive and less-redundant feature sets (Method 1 5.3; Supplementary appendix 2). Figure 7C shows the predictive power and minimum supervised distance of subsets of features refined by the feature selection approach. We can see that as the number of features reduces, the predictive performance drops, and the feature redundancy reduces (as indicated by an increasing minimum supervised distance). The figure shows that when using 90 features, the model can achieve good predictive performance (AUROC = 0.8845) and the minimum supervised distance within the features is high (0.9301). Figure 7D shows the summary plot of the top 10 features in the 5-year mortality prediction model using the selected 90 features. Since there is less redundancy in the selected features, we mitigate the issue of redundant features splitting credit. It allows us better to explore the effect of important risk predictors on mortality. In our low redundancy model, arm circumference is selected to represent the weight-related features and still receives high importance. Furthermore, we find that “requiring special healthcare equipment”, one of the top 10 features in the model trained on all features, is removed from the feature list because it is redundant with “general health condition”. In summary, our supervised distance-based feature selection method helps remove the redundant features and select the predictive and less-redundant feature set.

### 2.7 Highly accurate and efficient interpretable mortality risk scores

A mortality risk score can help individuals monitor their health status, help clinicians stratify high-risk patients, and help public health organizations guide policy. Most prior mortality risk scores are built with linear models, such as logistic regression, and linear hazard model [12, 18]. However, compared with traditional models, tree-based models achieve higher predictive performance, which can stratify patients better than linear models (Supplementary Table 1). Besides prediction performance, we also need to consider the feature collection cost. There is a tradeoff between collecting less features (which is less costly) and the model’s performance (cost-vs-accuracy tradeoffs). Moreover, the cost of features is different for different users. For example, blood test features are easily collected by clinicians, but for the public, questionnaire features and examination features are more feasible to obtain at home. Furthermore, the users may want to know which features contribute more to the risk score beside the risk score itself. To address these problems, we build interpretable tree-based mortality risk scores with different cost-vs-accuracy tradeoff and different types of features for the general public (demographic, examination, and questionnaire features) and medical professionals to use (demographic, laboratory features and features from common test panels) (Method 6; Supplementary appendix 2). Compared with previous mortality risk scores, ours are more interpretable, more accurate, applicable to more users, and flexible with different cost-vs-accuracy tradeoffs.

#### IMPACT develops highly accurate and efficient 5-year mortality risk scores

The predicted probability of IMPACT models can be directly used as mortality risk scores (IMPACT risk scores). We did a temporal validation of the risk scores by assessing their performances in the samples collected in NHANES 2009-2014. For comparison, we train linear and tree-based Cox proportional hazard models widely used in previous work. (Method 1 6.1) To find less costly but nearly as accurate models, we select the features using recursive feature elimination (RFE; Method 1 6.2). Moreover, we compare IMPACT risk scores with Intermountain sex-specific risk scores [18] ^10^ (Method 1 6.3). The models are evaluated on different gender groups.

In Figure 8A and B, we show the AUROC of the 5-year mortality risk scores of female samples (See Supplementary Figure 4 for male results) in the test set and the temporal validation set. We can see that the IMPACT model with only 20 features obtains an AUROC of 0.8971, which is almost as same as the performance of the model using all features (AUROC = 0.9030), and using fewer than 20 features leads to a dramatic accuracy drop. Figure 8A and B also show that IMPACT models achieve better performance than linear and tree-based Cox proportional hazard models. Furthermore, we can see that IMPACT risk score using the laboratory features (AUROC = 0.8881) and the risk score using the questionnaire and examination features (AUROC = 0.8835) both get acceptable predictive performance. The IMPACT risk score using the features from common test panels can achieve higher AUROCs than the intermountain risk score, which uses CBC and BMP panels features. With the models trained using different numbers of features, users can measure more risk predictors to use more accurate mortality prediction models. Figure 8B shows that the performance of our models only drops a little on the temporal validation set, which can indicate that our risk scores have generalization ability to some extent. The selected top 20 features and features from CBC and BMP panels are listed in Supplementary Table 2. In summary, we build IMPACT risk scores that are applicable to professional and non-professional individuals with different cost-vs-accuracy tradeoffs.

**Figure 8:**
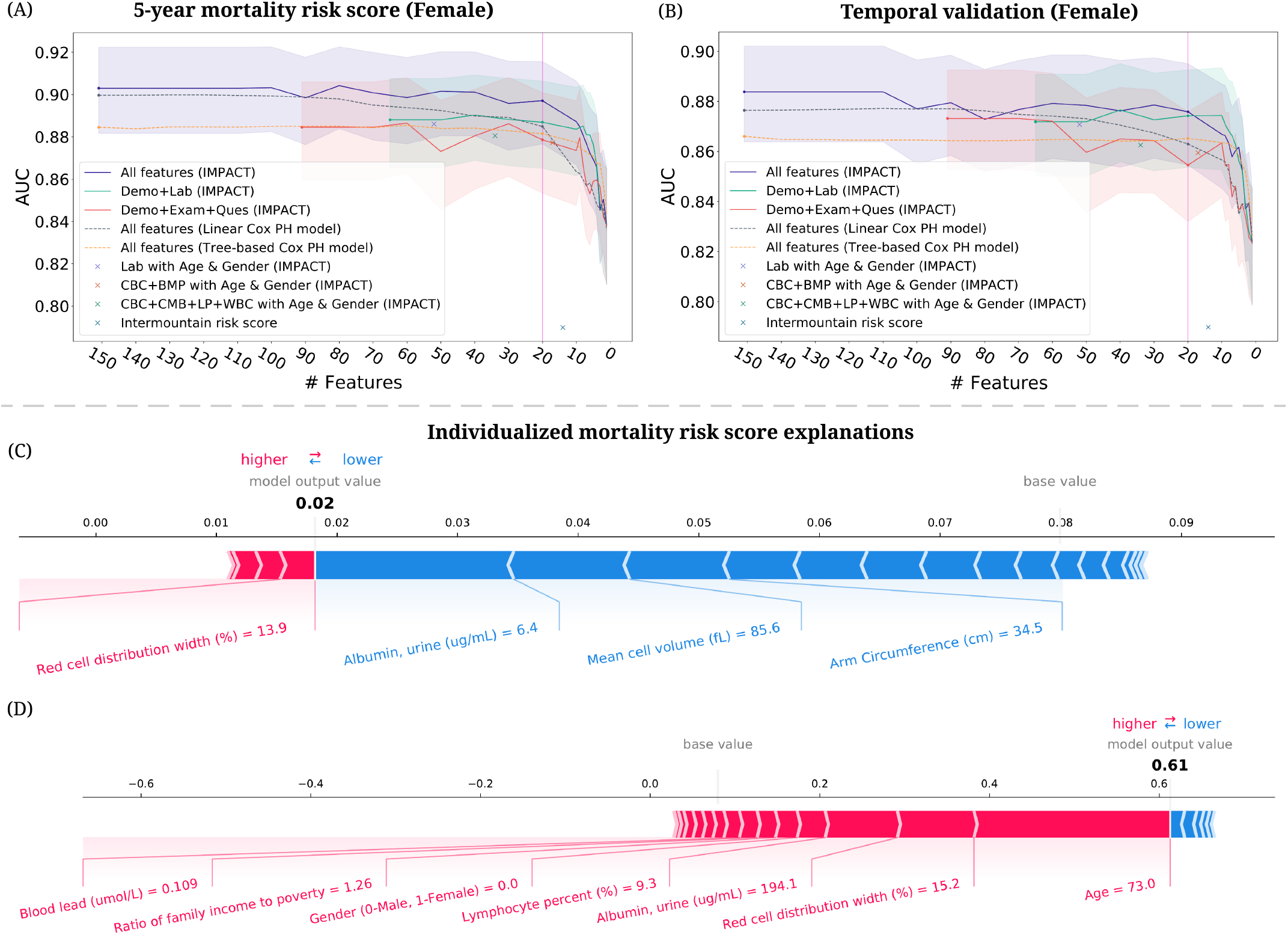
Developing highly accurate and efficient interpretable 5-year mortality risk scores. (A)–(B) The AUC of the models using different feature sets after recursive feature elimination. Lines are mean performance over 1000 random train/test splits, and shaded bands are 95 percent normal confidence intervals. (A) The AUC of the models tested on the female group in the test set of NHANES 1999-2008. (B) The AUC of the models testing on the female group in the temporal validation set (NHANES 2009-2014). (C)–(D) IMPACT can analyze individualized mortality risk scores. (C) The individualized explanation for an individual who is alive after 5 years. The output value is the risk score for that individual. The base value is the mean risk score, i.e., the score that would be predicted if we did not know any features for the current output. The features in red increase mortality risk, and those in blue decrease it. (D) The individualized explanation for a sample who is deceased after 5 years.

#### IMPACT exposes individualized mortality risk score explanations

TreeExplainer can help researchers analyze the prediction for each individual and illustrate each features’ contribution to the mortality risk score. We explain the mortality prediction model in terms of its probability predictions (risk scores) Method 1 6.1. Figure 8C,D shows individualized explanations for two individuals from the model using the top 20 features. The first individual (Figure 8C) was alive after 5 years. From the figure, we observe that IMPACT predicted that the individual’s 5-year mortality risk score was 0.02, lower than the average predicted risk (i.e., base value). There are features that increase mortality risk, such as red cell distribution width, and features that decrease mortality risk, such as urine albumin level. For this individual, the features that drive down mortality risk outweigh those that increase it. The second individual (Figure 8D) was deceased after 5 years, and the model’s predicted mortality probability is 0.61, much higher than the average predicted risk. The top three features that increase this individual’s risk are high age, high red cell distribution width, and high urine albumin concentration. These individualized explanations can help individuals understand their health status, adjust their lifestyle, and help doctors give personalized treatment and implement precision medicines.

## 3 Discussion

To our knowledge, IMPACT is the first study that combines high-accuracy complex ML models and state-of-the-art local explanation methods to do a systematic study of all-cause mortality. In epidemiology, high accuracy is important, but it is not enough; instead, explaining models to humans is essential for drawing epidemiological hypotheses [56, 57]. IMPACT’s combination of accuracy and explanation aims to optimize accuracy while also gaining insight into complex interrelations between mortality and individual’s features.

Using 151 features in NHANES 1999-2014, we build tree-based mortality prediction models and explore the effect of those features on mortality for different follow-up times and age groups. Importantly, we demonstrate the value and significance of explaining complex ML prognostic models. IMPACT allows us to capture both non-linear effects and interaction effects that are difficult to uncover with linear models. These results help us verify well-studied findings (e.g. the relationship of red cell distribution width and albumin with mortality) as well as identify new ones (e.g. the important risk predictors arm circumference, platelet count and serum chloride, and the complex interactions of the features). One pitfall to inferring relationships between determinants and an outcome are relationships between the determinants themselves (redundancy). To address this, we proposed a supervised distance and feature selection approach which we utilize to select the minimally redundant feature sets. Lastly, we build explainable mortality risk scores for both the general public and medical professionals with different tradeoffs between feature collection cost and the model’s performance. These scores can help individuals improve self-awareness of their health status and help clinicians identify patients with high mortality risk to target with specific interventions. In the paper, we only present a small part of our findings. All our results and risk scores are available in an interactive website^11^ where the associations and interaction can be explored in detail to generate new research hypotheses.

The present study shows a negative relationship between arm circumference and mortality. Our clustering method groups arm circumference with BMI and other weight-related features, indicating that these features share information about mortality. Several prior studies have found a U-shaped association between BMI and mortality, where very low or very high BMI is associated with significantly greater mortality risk [17, 2]. This U-shaped relationship may be the result of compound effects from body fat and fat-free mass. As upper arm circumference is an indicator of fat-free mass [64, 2], it may be the case that fat-free mass is driving the inverse correlation between arm circumference and mortality risk. Larger arm circumference is expected to be associated with greater muscle mass, while smaller arm circumference may reflect muscle deterioration along with diminished nutritional status or malnutrition [45, 59]. The importance of arm circumference in IMPACT is consistent with previous studies, which show that low arm circumference was more effective than low BMI in predicting follow-up mortality risk in older people [58, 45, 53].

One limitation of IMPACT is that the relationships and interactions detected by our model cannot be claimed to be causal. This is a common problem in epidemiological studies using observational data. The purpose of this study is not to address causality, but rather to do a systematic study of mortality associations with the NHANES population. In particular, one of the primary obstacles to capturing causal effects with observational data and predictive models are confounding variables. In order to condition on confounders (and potential surrogate confounders), it is often desirable to include as many features as possible in the model [46]. Conversely, we may want to remove colliders and mediators that skew the real effect of treatment features of interest. Our solution to redundancy, supervised distance, can potentially help narrow down related features for which domain experts can identify colliders, mediators, and confounders. This is a potential future research question which takes a step in the direction of making explanations from complex models causal.

As another limitation, our study is performed on NHANES 1999-2014, which is designed to assess the health status of participants in the United States. The conclusions and discoveries can generalize to other populations only when the distribution of variables and mortality rates are similar to those in the U.S. as a whole. Further external validation of our mortality models on datasets from non-U.S. populations should be undertaken to increase the generalizability of these findings.

Over the past several years, a variety of ML approaches have been applied in the field of aging research to develop “clocks” that are capable of predicting chronological age of an individual based on different phenotypic features [63]. The most common of these are the epigenetic clocks which have identified patterns of methylation on DNA that change with age and can be used to predict chronological age with high accuracy across a variety of different species and tissue types [20, 37]. Other clocks based on gene expression, metabolites, facial features, telomere length, etc. have also been described [60]. Efforts have also been made to use these clocks to predict an individual’s biological age, which may differ from their chronological age if they are aging more rapidly or slowly than the general population. Such “biological aging clocks” are expected to reflect the underlying health status of the individual and be useful for predicting future health outcomes and mortality. Although we have not yet attempted to validate IMPACT as a tool for assessing biological age, those individuals with significantly lower IMPACT mortality risk than expected for their chronological age would be predicted to have a lower biological age and vice-versa. Because IMPACT is trained to predict all-cause mortality rather than fit to chronological age, it will be of interest to determine how IMPACT compares to these various clocks in predictive capacity, particularly if done for the same cohort of individuals.

Prognosis research using complex ML models will likely increase over the coming years as ML techniques continue to rapidly develop. However, “black box” ML models that predict without explaining, are difficult for clinicians to trust and hard to extract meaningful information from. Therefore, the combination of complex ML models and ‘explainable artificial intelligence’ (XAI) is necessary and urgent. IMPACT takes a significant step towards XAI for mortality prediction. This study’s improvement in predictive accuracy and explanation of complex ML models warrants further exploration for other epidemiological outcomes.

## Method

### Method 1 Data collection and processing

The National Health and Nutrition Examination Survey (NHANES) from the National Center for Health Statistics (NCHS) ^1^ conducts interviews and physical examinations to assess the health and nutrition data for all ages in the United States. The interviews include demographic, socioeconomic, dietary, and health-related questions. The examinations include medical, dental, physiological measurements, and laboratory tests administered by highly trained medical personnel. Since 1999, data were collected and released at 2-year intervals. Each year NHANES examines a nationally representative sample of roughly 5,000 individuals across the Unites States. In this study, we include NHANES data sampled between 1999 and 2014. All-cause mortality is ascertained by a linked NHANES mortality file that provides follow-up mortality data from the date of survey participation through December 31, 2015.

Our study includes samples with known mortality status who participated in NHANES 1999-2014 (n = 47, 261). We include all demographic, laboratory, examination, and questionnaire features that could be automatically matched across different NHANES cycles. We exclude variables that are missing for more than 50% of the participants and highly correlated features with correlations greater than 0.98; after filtering and one-hot encoding, 151 features after one-hot encoding remain (Supplementary appendix 2). We impute missing data using MissForest [49], a nonparametric random forest-based multiple imputation method for mixed-type data, with seven iterations. We predict all-cause mortality for two broad categories: (1) follow-up times of 1-year, 3-year, and 5-year and (2) age groups of <40, 40-65, 65-80, and 80 years old. For different follow-up times, we remove samples with unconfirmed mortality status. For different age groups, we predict 5-year mortality. The demographic characteristics and sample size of the data for different tasks are shown in Reference Table 1.

### Method 2 Predictive modeling

To model mortality, we use gradient boosted trees (GBTs). GBTs are nonparametric methods composed of iteratively trained decision trees. The final ensemble of trees captures non-linearity and interactions between predictors. The dataset is randomly divided into training (80%) and testing (20%) sets. We use the implementation XGBoost [4] ^12^ with a learning rate set to 0.002, subsample ratio set to 0.5 and 10,000 trees of max depth 3. For comparison, we also train logistic regression models and deep neural networks. For logistic regression, we use L2 regularization. The L2 regularization weight was set to 100. For neural networks, we use a single layer with 1,000 nodes, and max iteration set to 1,000. The hyperparameters specified above are chosen by GridSearch and 5-fold cross validation. Other hyperparameter values are left at their default values. Models’ performance is measured with the area under the receiver operator characteristic curve (AUROC). We bootstrap the test set to assess the statistical significance of the difference in AUC for pairs of models. Specifically, we resample with replacement from the test set 1,000 times and compare the models’ performance on resampled test sets. We report a p-value which is the percentage of time that logistic regression or the neural network’s performance is better than or equal to gradient boosted trees, divided by the number of resampled test sets. All models are built using the Scikit-learn package in Python 3.7.

### Method 3 Model interpretation

To explain the GBT models, we utilize TreeExplainer [28], which provides a local explanation of the impact of input features on individual predictions. Specifically, TreeExplainer calculates exact SHAP [27] (SHapley Additive exPlanations) values for tree-based models. When explaining the mortality prediction models, we randomly select 10,000 background samples from the training set and 5,000 foreground samples from the test set.

#### Method 1 3.1 SHAP (SHapley Additive exPlanation) values

SHAP (SHapley Additive exPlanation) values attribute to each feature the change in the expected model prediction when conditioning on that feature. The change of the model’s prediction when the feature is masked is recorded across all possible subsets of features, yielding an average change in prediction resulting from the inclusion of a feature in the model:

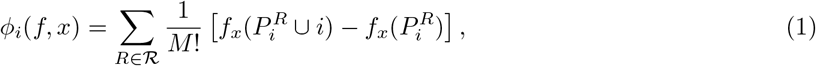

where ϕ_*i*_ is the feature attribution (SHAP value) of feature *i* in model *f* for data point *x*, ℛ is the set of all feature permutations, 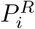 is the set of all features before i in the ordering *R, M* is the number of input features, and *f*_*x*_ is an estimate of the conditional expectation of the model’s prediction: *f*_*x*_(*S*) ≈ E[f(x) | x_S_] where *x*_*S*_ is the set of observed features.

SHAP values which guarantee a set of desirable theoretical properties, including additivity and consistency. Additivity states that when approximating the original model *f* for a specific input x, the SHAP values sum up to the output *f*(*x*):

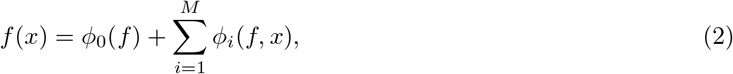

The sum of feature attributions (SHAP values) matches the original model output f(x), where ϕ_0_(*f*) = E[f(z)] = f_x_(Ø). Consistency states that if a model changes so that some feature’s contribution increases or stays the same regardless of the other inputs, that input’s attribution should not decrease. Therefore, SHAP values are consistent and accurate calculations of each feature’s contribution to the model’s prediction.

#### Method 1 3.2 SHAP interaction values and main effects

The SHAP interaction effects is based on the Shapley interaction index from game theory. While standard feature attribution results in a vector of values, one for each feature, attributions based on the Shapley interaction index result in a matrix of feature attributions. The main effects are on the diagonal and the interaction effects on the off-diagonal. The SHAP interaction values are defined as:

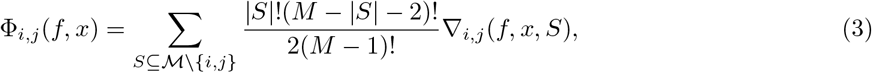

when *i* ≠ *j*, and

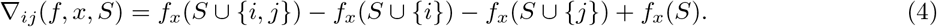

where ℳ is the set of all *M* input features. In Equation 3 the SHAP interaction value between feature *i* and feature *j* is split equally between each feature so Φ_*i,f*_ (*f, x*) = Φ_*j,i*_(*f, x*) and the total interaction effect is Φ_*i,f*_ (*f, x*) + Φ_*j,i*_(*f, x*).

The main effects for a prediction can then be defined as the difference between the SHAP values and the off-diagonal SHAP interaction values for a feature:

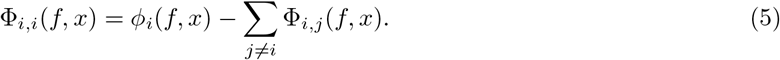

#### Method 1 3.3 Partial dependence plots and additional perspective to reference interval

We use the partial dependence plots to show the change in mortality risk for all values of a laboratory feature. The partial dependence plot shows the marginal effect one features have on the predicted outcome of a machine learning model. The relative mortality risk is defined as the average value of the model predicted probability when we fix a specific feature to a given value divided by the average value of the model predicted probability. The relative risk percentage is the maximum relative risk for the values within the reference interval divided by the maximum relative risk for all values of a laboratory feature. High relative risk percentage indicates that the values within the reference interval have a relatively high mortality risk. The partial dependence plots of selected laboratory feature values on 1-, 3-, and 10-year mortality risk are shown in Supplementary Figure 5.

### Method 4 Model interpretation plots

In this section we describe a number of plotting types for model explanation visualization.

#### SHAP value, SHAP main effect value and SHAP interaction value plots

In SHAP value/SHAP main effect value/SHAP interaction value plots, every point corresponds to a single sample where the x-axis is the value of the feature and the y-axis is the SHAP value/SHAP main effect value/SHAP interaction value. The coloring of the points often denotes the value of a separate feature.

#### Summary plot

Summary plots show the feature attributions (SHAP values) for many samples and multiple features in order of global feature importance (the mean absolute SHAP values). Summary plots stack multiple subplots plots for each feature. For the feature plots, every point corresponds to a single sample where the x-axis is the feature attribution value and the y-axis is vertical dispersion representing the frequency of samples with a particular feature attribution value. Finally, the color of each point represents the normalized feature value, with red representing a high value and blue representing a low one. Intermediary feature values are interpolations between red and blue.

#### Individualized explanation plot

Individualized explanation plot shows the feature attributions (SHAP values) for an individual in terms of how they drive the model’s prediction for the individual away from the average model prediction across the baseline distribution. The width of the bars indicate the SHAP value with red indicating a positive affect and blue indicating a negative one. The features corresponding to the largest bars are below with their actual values for the individual.

### Method 5 Supervised distance

#### Method 1 5.1 Supervised distance and hierarchical clustering

Supervised distance can accurately measure feature redundancy based on a specific prediction task. As Supplementary Figure 2 shows, to calculate the supervised distance between feature *i* and feature *j*, we firstly train a uni-variate GBTs model to predict the label (e.g. 5-year mortality in our study) using feature *i*. Then, we can obtain the *Prediction*_*i*_ which is the output of the fitted uni-variate GBTs. Next, we fit another uni-variate GBTs to predict *Prediction*_*i*_ using feature *j*. We define the output of the new GBTs as 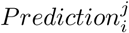. All hyperparameter values of the uni-variate GBTs are set to their default values. Following the same above steps, we can obtain 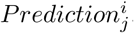. The supervised distance between feature *i* and feature *j* (supervised distance(*i,j*)) is defined as:

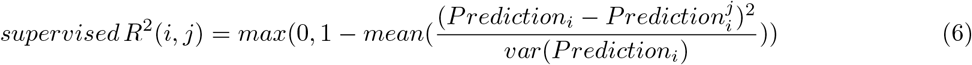

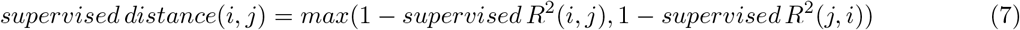

where *var*(*x*) is the variance of the vector *x, mean*(*x*) is the average of the vector *x*. Supervised distance is scaled roughly between 0 and 1, where 0 distance means the features perfectly redundant and 1 means they are completely independent.

To explore the redundant feature groups, we hierarchically cluster all features according to the supervised distance. Specifically, we use complete linkage hierarchical clustering which merges in each step the two clusters whose merger has the smallest diameter. The hierarchical clustering tree is shown in Supplementary Figure 3.

#### Method 1 5.2 Redundant feature groups experiments training details

##### Reducing redundancy model

To identify the most representative feature in a redundant feature group, we train GBTs using one feature in the redundancy group and all features outside the group for 5-year mortality prediction. Then we compare the feature importance ranking of the redundant features by calculating the mean absolute SHAP values using TreeExplainer. The hyperparameters of the GBTs are chosen by GridSearch and 5-fold cross validation. The max depth is selected from {1, 3, 5, 7, 9} and the subsample ratio is selected from {0.2, 0.5, 0.8, 1.0}. Other hyperparameter values are left at their default values.

##### Single feature model

We further analyze the predictive power of the redundant features by fitting 5-year a GBTs using one feature in the redundant feature group. Specifically, we use one feature in the redundant feature group and two important confounders, age and gender, to train a GBTs for 5-year mortality prediction. All hyperparameter values are set to their default values. We compare the AUCs of the models. We bootstrap the test set for 1,000 times and compare the models’ performance on resampled test sets. The averages of the AUCs are reported.

### Method 1 5.3 Supervised distance-based feature selection

We propose a supervised distance-based feature selection method to select predictive and less-redundant feature sets. The workflow of our feature selection method is shown in Supplementary Figure 2. The dataset is randomly divided into training (80%) and testing (20%) sets. Firstly, we fit a GBTs for 5-year mortality prediction on all features using the training set and rank the features by mean absolute SHAP values from TreeExplainer. The hyperparameters of the GBTs are chosen by GridSearch and 5-fold cross validation. The max depth is selected from {1, 3, 5, 7, 9} and the subsample ratio is selected from {0.2, 0.5, 0.8, 1.0}. The max number of trees is set to 1000. We use 20% of the training samples as validation set for early stopping. The number of early stopping rounds is set to 100. Since age and gender are important confounders, we would like to keep them in the selected feature set. Therefore, we cluster features except age and gender into a specific number of groups using supervised distances-based hierarchical clustering and select the most important feature in each cluster. Then, we add age and gender to the selected feature set and re-fit the model. Next, we rerun the clustering using the new feature set except age and gender. This process is repeated until all remained features cluster to a single group. In every iteration, we remove 5 features. The models are evaluated on the testing set with bootstrapping for 1,000 times. We report the average of the AUCs and the minimum supervised distance within the selected feature sets. The selected features in each iteration are listed in Supplementary Appendix 1.

### Method 6 5-year mortality risk scores

#### Method 1 6.1 Mortality risk scores training details

IMPACT mortality risk scores is defined as the prediction of the 5-year mortality prediction models. For comparison, we train linear ^13^ and gradient boosted tree-based Cox proportional hazard models ^14^. We did a temporal validation of the risk scores by assessing their performances in the samples collected in 2009-2014 (N = 7, 034). Specifically, the samples collected in 1999-2008 (N = 28, 820) are randomly divided into training (80%) and testing (20%) sets. To compare with Intermountain gender-specific risk scores, we evaluate the models on different gender groups. The models are trained on the whole training set and evaluate on different gender groups in the testing set. Furthermore, considering the different feature collection cost for general public and medical professionals, we build the risk scores using different feature sets. For general public, the models are trained on all demographics, questionnaire features and examination features that are accessible at home for general public, For medical professionals, the models are trained on all demographics and laboratory features. All trained models are evaluated on different gender groups of the samples collected in 2009-2014 for temporal validation.

The hyperparameters are chosen by GridSearch and 5-fold cross validation. For XGBoost 5-year mortality prediction models, the max depth is selected from {1, 3, 5, 7, 9} and the subsample ratio is selected from {0.2, 0.5, 0.8, 1.0}. The max number of trees is set to 1000. We use 20% of the training samples as validation set for early stopping. The number of early stopping rounds is set to 100. For linear Cox proportional hazard models, the regularization parameter *α* is selected from {0.01, 0.1, 1, 10, 100}. For tree-based Cox proportional hazard models, the max depth is selected from {1, 3, 5, 7, 9} and the subsample ratio is selected from {0.2, 0.5, 0.8, 1.0}. Other hyperparameter values are left at their default values.

We explain the mortality prediction model in terms of its probability predictions. Specifically, we rescales the SHAP values (in the log-odds space) to be in the probability space directly. The rescaled SHAP values now sum to the probability output of the model.

#### Method 1 6.2 Recursive feature elimination

Recursive feature elimination works by searching for a subset of features by starting with all features in the training dataset and successfully removing features until the desired number of feature remain. Firstly, we train a model on the full dataset with all features. Then we rank features by importance (mean absolute SHAP values) and remove the least important features. Another model is trained on the resulting feature set, and the process iterates until only the desired number of features are left. Starting from 151 features, we remove 6 features at the first iteration Then, We remove 5 features in each iteration until only one feature left. We bootstrap the test set and assess the predictive performance. Specifically, we resample with replacement from the test set 1,000 times and report the average and the 95% confidence interval of the AUCs. The selected features in each iteration are listed in Supplementary appendix 2.

#### Method 1 6.3 Intermountain mortality risk scores and exhaustive feature selection

Intermountain mortality risk scores [18] are built using complete blood count and basic metabolic profile. Specifically, 13 laboratory features are used to predict 30 days, 1-year and 5-year mortality. Logistic regression was used to model the risk prediction equations with adjustment for age and sex. Dummy variables modeled each category, with the referent defined as the lowest risk group (except for age categories: 18-29, 30-39, 40-49 [referent], 50-59, 60-69, 70-79, and ≥80 years). A scalar score value was derived for each variable category by multiplying its *β*-coefficient by 3 and rounding to the nearest integer (referent value = zero). Each individual’s risk score became the sum of the score values based on his or her individual data.

We implement exhaustive search to select features of Intermountain risk scores. The number of features ranges from 1 to 14 (including age). Given the number of features, we search all possible feature combinations. The risk score becomes the sum of the score values of the selected features. The 5-year mortality risk scores are evaluated on the training set. We select the feature combination that achieve the highest AUC on the training set. Then, the risk scores of the selected feature combinations are evaluated on the testing set with bootstrapping for 1,000 times.

## Data Availability

Data is available from The National Health and Nutrition Examination Survey (NHANES - https://www.cdc.gov/nchs/nhanes/).

https://github.com/qiuweipku/IMPACT

## Data availability

The data for all experiments and figures in the paper are publicly available. A downloadable version of the dataset is available at https://github.com/qiuweipku/IMPACT.

## Code availability

The code for our study is available at https://github.com/qiuweipku/IMPACT. The code for our interactive website is available at https://github.com/qiuweipku/impact-website.

## Acknowledges

This work was funded by National Science Foundation [DBI-1759487, DBI-1552309, DBI-1355899, DGE-1762114]; National Institutes of Health [R35 GM 128638, R01 NIA AG 061132 and P30 AG 013280].

## Footnotes

1 https://qiuweipku.github.io/IMPACT

1 http://www.cdc.gov/nchs/nhanes.htm

3 https://www.webmd.com/hepatitis/ggt-test

4 https://medlineplus.gov/ency/article/003544.htm

5 https://www.ucsfhealth.org/medical-tests/blood-differential-test

6 https://www.ucsfhealth.org/medical-tests/blood-differential-test

7 https://www.mayocliniclabs.com/test-catalog/Clinical+and+Interpretive/610525

8 https://www.ucsfhealth.org/medical-tests/lead-levels—blood

9 https://cllsociety.org/toolbox/normal-lab-values/

10 https://intermountainhealthcare.org/IMRS/

11 https://qiuweipku.github.io/IMPACT

1 http://www.cdc.gov/nchs/nhanes.htm

12 https://xgboost.readthedocs.io/en/latest/python/index.html

13 https://scikit-survival.readthedocs.io/en/latest/api/generated/sksurv.linearmodel.CoxPHSurvivalAnalysis.html

14 https://scikit-survival.readthedocs.io/en/latest/api/generated/sksurv.ensemble.GradientBoostingSurvivalAnalysis.html

