## Supplementary Appendix 1 for "Interpretable machine learning prediction of all-cause mortality"

### 1 Discoveries for mortality prediction using different age groups.

***IMPACT identifies important features for mortality prediction in different age groups.*** Supplementary Figure 1A shows the top 20 most important features and relative importance in 5-year mortality prediction models using different age groups ($<$40, 40-65, 65-80 and $\geq$80). Some features become more important for older subpopulations, such as alanine aminotransferase (ALT), the fifth most important feature in the model using samples over 80 years. Supplementary Figure 1E shows the main effect of ALT for age$\geq$80, which shows the negative relationship between ALT and 5-year mortality. Moreover, some features are less important for older subpopulations than younger ones. One example is uric acid level, the sixth most important feature in the age$<$40 model and the 59th most important in the age$\geq$80 model. Supplementary Figure 1B plot the main effect and SHAP value of uric acid in the age$<$40 model, showing that low uric acid levels increase mortality risk prediction. However, in the age 40-65 model, higher uric acid is associated with higher mortality risk (Supplementary Figure 1D). Previous work shows that low uric acid in blood serum can injure the endothelium and induce oxidative stress-related disease [4, 6], and that hyperuricemia (high uric acid) is associated with various adverse health outcomes, including hypertension, stroke, cardiovascular disease and cancer [1, 2, 3, 5]. The numerous downstream effects of high uric acid and low uric acid might explain the different relationship between uric acid and mortality in different age groups. Moreover, the reference range of uric acid differs for males and females (2.4-6.0 mg/dL for females and 3.4-7.0 mg/dL for males). This difference is shown in Figure 1C, where women have lower uric acid, which can increase mortality risk.

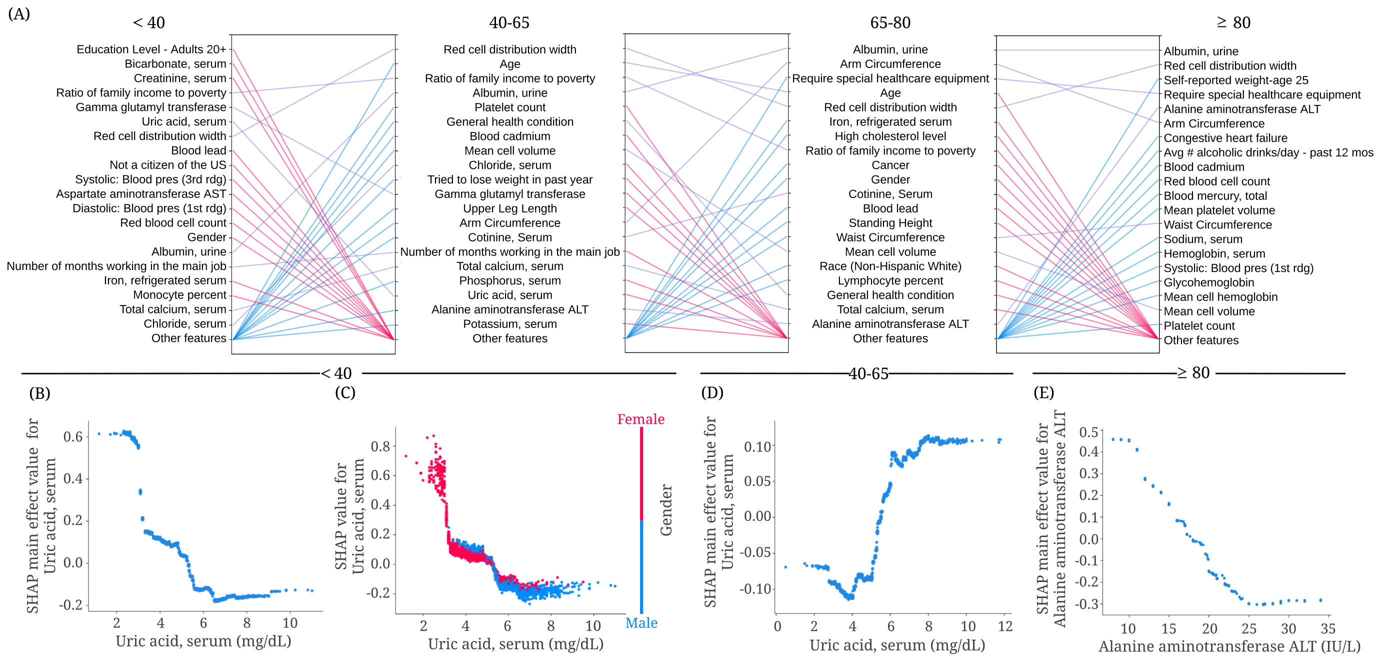


Supplementary Figure 1: **Identifying important discoveries for mortality prediction in different age groups.** (A) Relative importance of input features in <40, 40-65, 65-80 and ≥80 age groups. For each model, the figure shows the 20 most impactful features on prediction (ranked from most to least important). The purple line indicates that the feature is in the top 20 features of two models. Blue and red lines indicate the feature is in the top 20 features of one model, but not in the top 20 features of the other. (B) The main effect of serum uric acid on 5-year mortality in the <40 age group. (C) The SHAP value of serum uric acid in the <40 age group 5-year mortality model. (D) The main effect of serum uric acid on 5-year mortality in the 40-65 age group. (E) The main effect of alanine aminotransferase on 5-year mortality in the ≥80 age group.

| Different follow-up times | | | | | |
|---|---|---|---|---|---|
| | | 1-year (n=47,261) | 3-year (n=41,434) | 5-year (n=35,854) | 10-year (n=21,986) |
| Number of deaths | | 577 (1.22%) | 1,897 (4.58%) | 3,074 (8.57%) | 5,295 (24.08%) |
| Age,years | | 46 (30-63) | 46 (30-64) | 46 (30-64) | 48 (30-67) |
| Sex | | | | | |
| | Male | 22,778 (48.20%) | 19,998 (48.26%) | 17,276 (48.18%) | 10,630 (48.35%) |
| | Female | 24,483 (51.80%) | 21,436 (51.74%) | 18,578 (51.82%) | 11,356 (51.65%) |
| Ethnicity | | | | | |
| | Mexican American | 8,947 (19.93%) | 8,164 (19.70%) | 7,543 (21.04%) | 4,844 (22.03%) |
| | Other Hispanic | 3,452 (7.03%) | 2,929 (7.07%) | 2,335 (6.51%) | 979 (4.45%) |
| | Non-Hispanic White | 21,428 (45.34%) | 18,990 (45.83%) | 17,081 (47.64%) | 10,921 (49.67%) |
| | Non-Hispanic Black | 10,039 (21.24%) | 8,821 (21.29%) | 7,337 (20.46%) | 4,353 (19.78%) |
| | Other Race | 3,395 (7.18%) | 2,530 (6.11%) | 1,558 (4.35%) | 889 (4.04%) |
| Different age groups (follow-up time = 5-year) | | | | | |
| | | Age $<$ 40 (n=14,227) | 40 $\leq$ Age $<$ 65 (n=12,871) | 65 $\leq$ Age $<$ 80 (n=7,457) | Age $\geq$ 80 (n=1,299) |
| Number of deaths | | 187 (1.31%) | 718 (5.58%) | 1,593 (21.36%) | 576 (44.34%) |
| Age,years | | 27 (21-33) | 51 (45-59) | 72 (68-75) | 80 (80-80) |
| Sex | | | | | |
| | Male | 6,629 (46.59%) | 6,315 (49.06%) | 3,710 (49.75%) | 622 (47.88%) |
| | Female | 7,598 (53.41%) | 6,556 (50.94%) | 3,747 (50.25%) | 677 (52.12%) |
| Ethnicity | | | | | |
| | Mexican American | 3,663 (25.75%) | 2,633 (20.46%) | 1,149 (15.41%) | 98 (7.54%) |
| | Other Hispanic | 993 (6.98%) | 918 (7.113%) | 362 (4.85%) | 62 (4.77%) |
| | Non-Hispanic White | 5,692 (40.01%) | 5,945 (46.19%) | 4,493 (60.25%) | 951 (73.21) |
| | Non-Hispanic Black | 3,143 (22.09%) | 2,808 (21.82%) | 1,250 (16.76%) | 136 (10.47) |
| | Other Race | 736 (5.17%) | 567 (4.41%) | 203 (2.72%) | 52 (4.00%) |

Supplementary Table 1: Population characteristics for the study cohorts. Data are median (IQR), or n/N (%).

| Importance Ranking | IMPACT-20 | IMPACT-20 (Demo+Lab) |
|---|---|---|
| 1 | Age | Age |
| 2 | Albumin, urine (ug/mL) | Blood lead (umol/L) |
| 3 | Arm Circumference (cm) | Albumin, urine (ug/mL) |
| 4 | Gender (0-Male, 1-Female) | Ratio of family income to poverty |
| 5 | Blood lead (umol/L) | Education Level - Adults 20+ |
| 6 | Ratio of family income to poverty | Red cell distribution width (%) |
| 7 | Albumin, serum (g/L) | Chloride, serum (mmol/L) |
| 8 | Red cell distribution width (%) | Blood cadmium (nmol/L) |
| 9 | Received Hepatitis B 3 dose series | Lymphocyte percent (%) |
| 10 | General health condition | Mean cell volume (fL) |
| 11 | Mean cell volume (fL) | Red blood cell count (million cells/uL) |
| 12 | Number of months working in the main job | Albumin, serum (g/L) |
| 13 | Self-reported greatest weight (pounds) | Creatinine, serum (umol/L) |
| 14 | Education Level - Adults 20+ | Cotinine, Serum (ng/mL) |
| 15 | Lymphocyte percent (%) | Platelet count (1000 cells/uL) |
| 16 | Require special healthcare equipment (0-No, 1-Yes) | Potassium, serum (mmol/L) |
| 17 | Chloride, serum (mmol/L) | Sodium, serum (mmol/L) |
| 18 | Blood cadmium (nmol/L) | Alanine aminotransferase ALT (IU/L) |
| 19 | Weight (kg) | Blood urea nitrogen (mmol/L) |
| 20 | Shortness of breath on stairs/inclines (0-No, 1-Yes) | Race (Non-Hispanic White) |

| Importance Ranking | IMPACT-20 (Demo+Exam+Ques) | IMPACT (CBC+BMP with age and gender) |
|---|---|---|
| 1 | Age | Age |
| 2 | Require special healthcare equipment (0-No, 1-Yes) | Red cell distribution width (%) |
| 3 | Arm Circumference (cm) | Mean cell volume (fL) |
| 4 | General health condition | Chloride, serum (mmol/L) |
| 5 | Education Level - Adults 20+ | Gender (0-Male, 1-Female) |
| 6 | Gender (0-Male, 1-Female) | Glucose, refrigerated serum (mmol/L) |
| 7 | Congestive heart failure (0-No, 1-Yes) | Red blood cell count (million cells/uL) |
| 8 | Ratio of family income to poverty | White blood cell count (1000 cells/uL) |
| 9 | Diastolic: Blood pres (2nd rdg) mm Hg | Potassium, serum (mmol/L) |
| 10 | Systolic: Blood pres (2nd rdg) mm Hg | Creatinine, serum (umol/L) |
| 11 | Avg # alcoholic drinks/day - past 12 mos | Platelet count (1000 cells/uL) |
| 12 | Cancer (0-No, 1-Yes) | Blood urea nitrogen (mmol/L) |
| 13 | Self-reported weight-age 25 (pounds) | Sodium, serum (mmol/L) |
| 14 | Number of months working in the main job | Hemoglobin, serum (g/dL) |
| 15 | Self-reported greatest weight (pounds) | Mean cell hemoglobin (pg) |
| 16 | Duration of longest job (months) | Total calcium, serum (mmol/L) |
| 17 | Smoked at least 100 cigarettes in life (0-No, 1-Yes) | Mean platelet volume (fL) |
| 18 | Shortness of breath on stairs/inclines (0-No, 1-Yes) | |
| 19 | 60 sec. pulse (30 sec. pulse * 2) | |
| 20 | Current self-reported height (inches) | |

Supplementary Table 2: The selected top 20 features of 5-year mortality risk scores using different feature types and the features included in CBC and BMP panels.

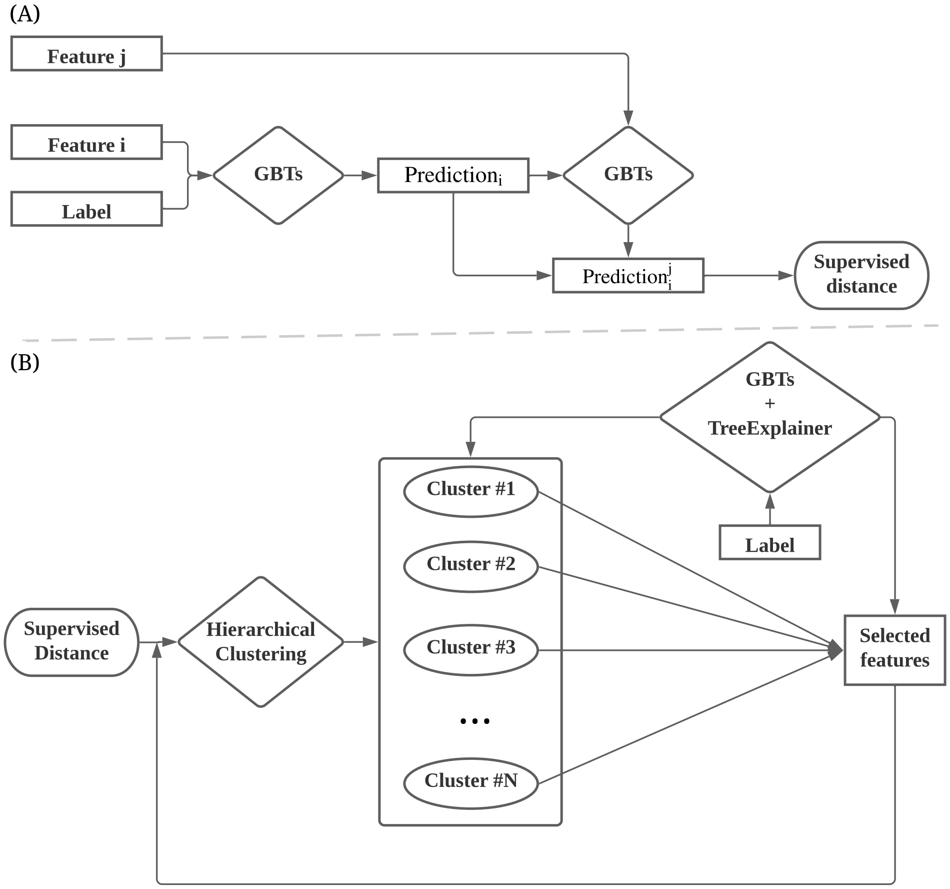


Supplementary Figure 2: (A) The workflow of supervised distance calculation. (B) The workflow of supervised-distance feature selection.

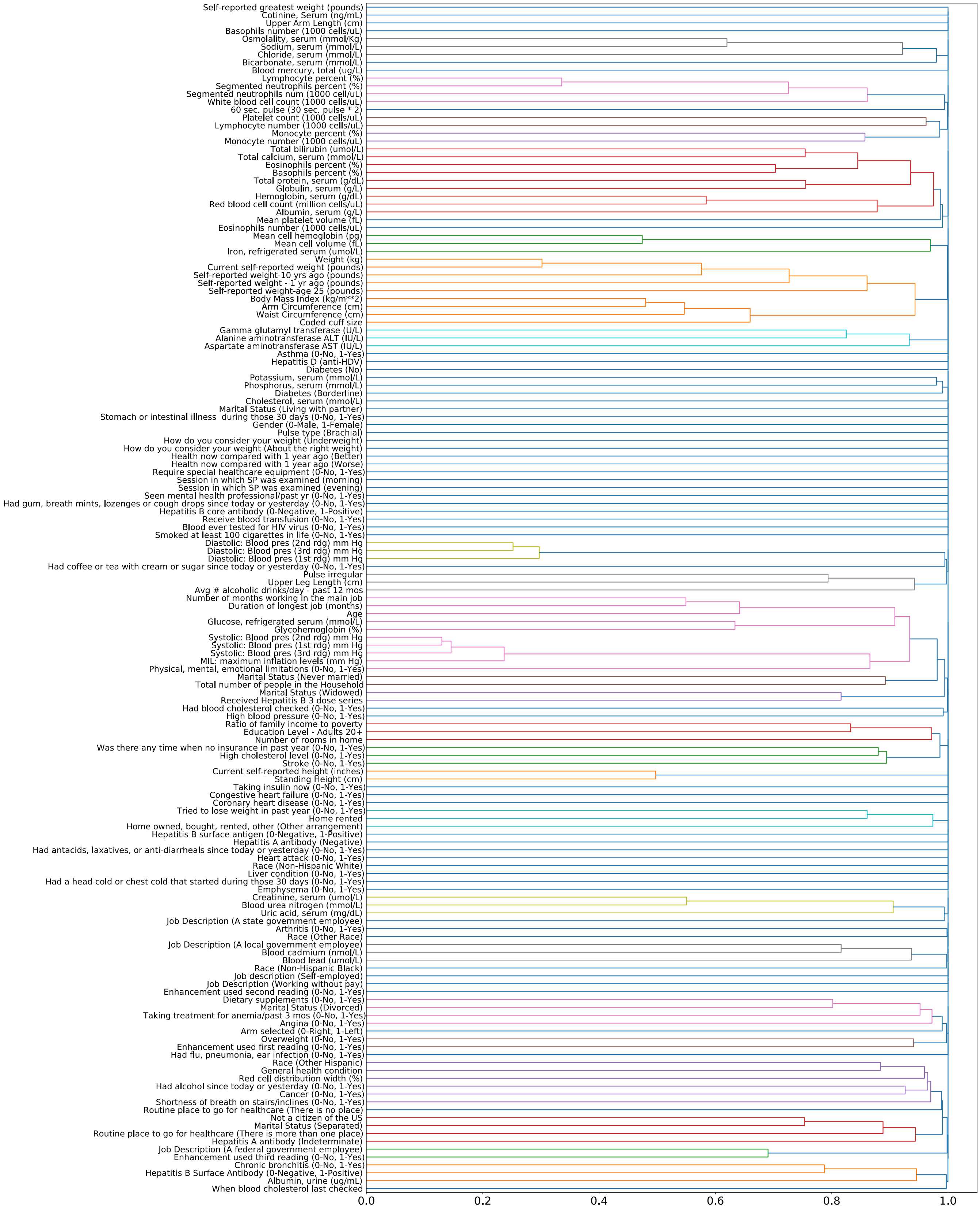


Supplementary Figure 3: The cluster tree of supervised distance based hierarchical clustering. The color threshold is set to 0.98.

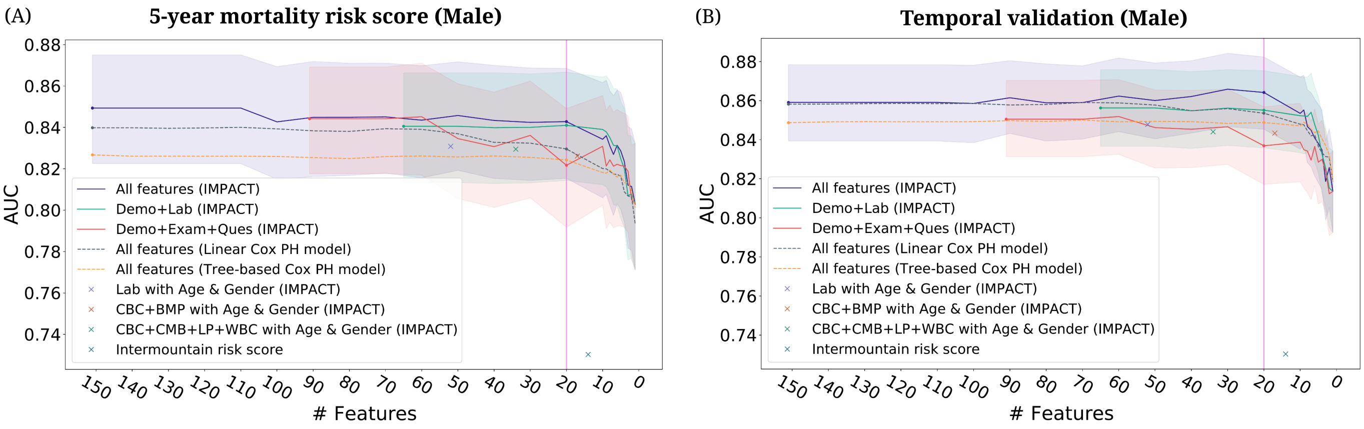


Supplementary Figure 4: (A)–(B) The AUC of the models using different feature sets after recursive feature elimination. Lines are mean performance over 1000 random train/test splits, and shaded bands are 95 percent normal confidence intervals. (A) The AUC of the models tested on the male group in the test set of NHANES 1999-2008. (B) The AUC of the models testing on the male group in the temporal validation set (NHANES 2009-2014).

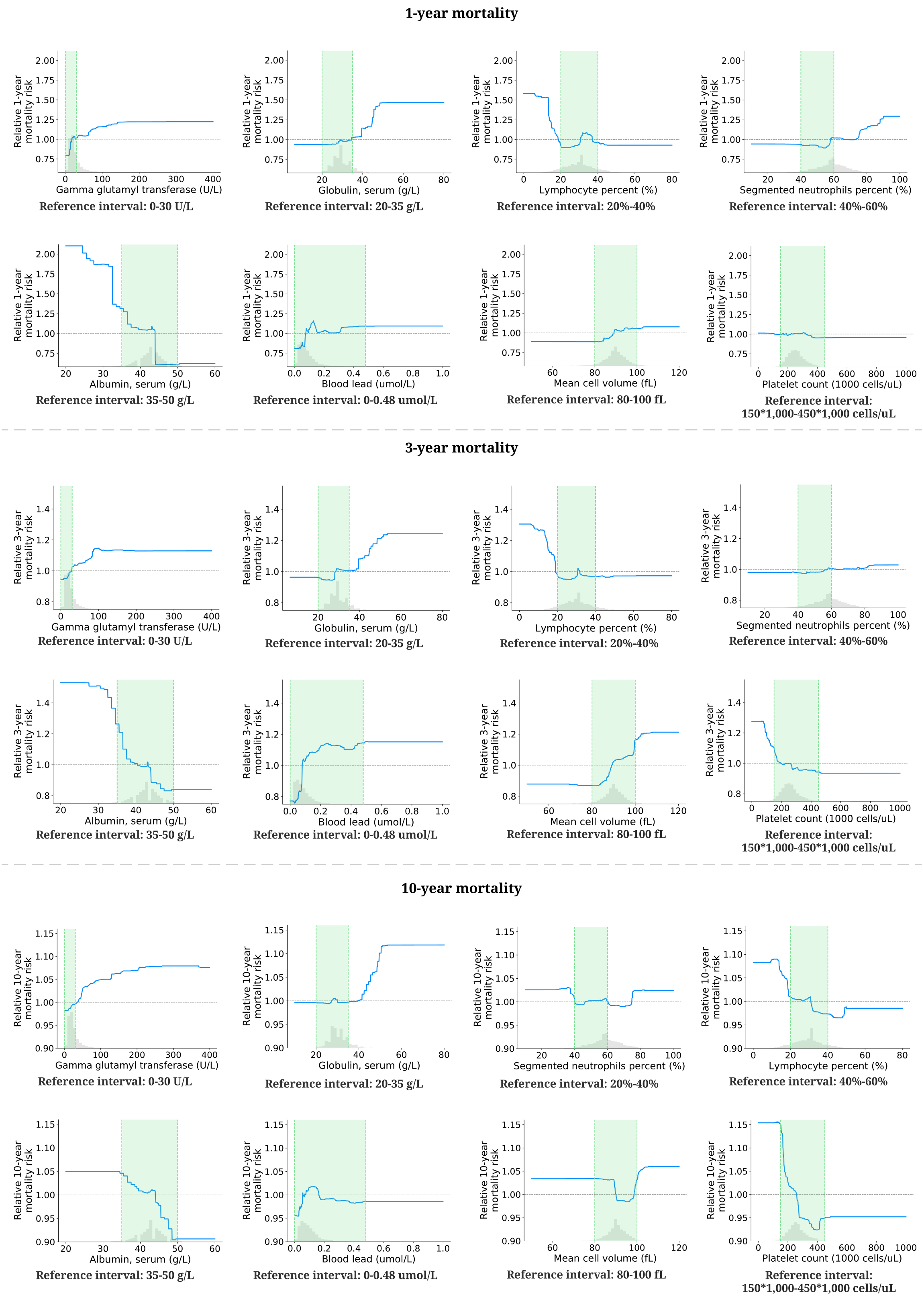


Supplementary Figure 5: **Effect of varying laboratory feature values on 1-, 3- and 10-year mortality risk.** These partial dependence plots show the change in relative 1-, 3- and 10-year mortality risk (Method 1 3.3) for all values of a given laboratory feature. The grey histograms on each plot show the distribution of values for that feature in the test set. The green shaded region shows the reference interval of each feature.
